# Immunomodulatory metabolites define long-term gut microbiome recovery after allogeneic HCT and associate with improved survival and reduced relapse related mortality

**DOI:** 10.64898/2026.03.26.26349381

**Authors:** Alix Schwarz, Tina Eismann, Tingting Zheng, Simon Holzinger, Alexander Denk, Sophia Goeldel, Mats Urban, Sascha Göttert, Mohsen Pourjam, Ilias Lagkouvardos, Klaus Neuhaus, Peter Herhaus, Mareike Verbeek, Romana R. Gerner, Matthias Fante, Andreas Hiergeist, André Gessner, Matthias Edinger, Wolfgang Herr, Karin Kleigrewe, Simon Heidegger, Klaus-Peter Janssen, Ernst Holler, Elisabeth Meedt, Melanie Schirmer, Florian Bassermann, Daniel Wolff, Hendrik Poeck, Daniela Weber, Erik Thiele Orberg

## Abstract

The intestinal microbiome influences immune recovery and long-term outcomes after allogeneic hematopoietic stem cell transplantation (allo-SCT). While reduced bacterial diversity and depletion of immunomodulatory microbial metabolites during peri-engraftment have been linked to acute graft-versus-host disease (aGvHD) and mortality, it remains unclear whether microbiome recovery after engraftment and immune reconstitution is better reflected by bacterial diversity or by microbial metabolic output. We aimed to define microbiome recovery in the late post-transplant period and test whether a metabolite-based biomarker improves the prediction of clinical outcomes, including overall survival (OS) and chronic (c) GvHD.

In this two-center longitudinal observational study, serial stool samples were collected from pre-transplant baseline to day +100 after allo-SCT in a discovery cohort (n = 20, Technical University Munich University Hospital (TUM)) and an independent validation cohort (n = 100, University Hospital Regensburg (UKR)). Gut microbiome composition was assessed by 16S rRNA gene amplicon sequencing, with metagenomic profiling in selected patients, and stool metabolites were quantified using targeted mass spectrometry. Patients were classified as *RECOVERY* or *NO RECOVERY* based on changes in bacterial richness between baseline and the post-transplant period. To capture microbial metabolic output, the previously established *Immune-Modulatory Metabolite Risk Index* (*IMM-RI*), comprising butyric, propionic, and isovaleric acids, desaminotyrosine and indole-3-carboxaldehyde, was adapted to the late posttransplant period (*IMM-RI post-TX*).

Bacterial alpha diversity frequently improved by day +100; however, this did not consistently indicate restoration of baseline community structure and was not paralleled by recovery of stool metabolite profiles. Accordingly, *RECOVERY* status showed a limited association with survival or transplant-related mortality (TRM). In contrast, *IMM-RI post-TX* low-risk identified patients with preserved butyrate-associated biosynthetic capacity and was significantly associated with improved OS in both cohorts (UKR: HR 0.2052, 95% CI 0.07703 – 0.5466, p < 0.0001). In the validation cohort, *IMM-RI post-*TX low-risk was significantly associated with reduced relapse related mortality. Interestingly, stool butyric-, propionic and valeric acid concentrations were increased in cGvHD of the skin, indicating context-dependent metabolite effects.

These findings suggest that metabolite profiling outperforms bacterial diversity for predicting outcomes after allo-SCT and support microbial metabolites as promising biomarkers for risk stratification and actionable candidates for precision microbiome interventions after allo-SCT.

**Data sharing statement:**

- For original data, please contact

**Key points:**

- Metabolomic profiling outperforms microbial taxonomy for predicting long-term clinical outcomes in allo-SCT recipients
- Immune-Modulatory Metabolite Risk Index post-transplant is significantly associated with overall survival, relapse rate and incidence of chronic GvHD

## INTRODUCTION

Allogeneic hematopoietic stem cell transplantation (allo-SCT) is a curative treatment for patients suffering from hematological malignancies.^1,2^ Despite the advent of new prophylactic strategies including post-transplant cyclophosphamide, a significant proportion of patients develop aGvHD, which occurs when co-transplanted donor-derived T cells attack recipients’ tissues. Depending mainly on the degree of HLA mismatch and the GvHD prophylaxis administered, the incidence of grade II-IV aGvHD ranges between 40% and 62%.^3^ About 30%-50% of patients become refractory to steroid treatment, and overall aGvHD accounts for approximately 20-30% of TRM.^4–7^ Other subsequent immunosuppressive therapies-lines are associated with increased infectious risks and a potential loss of the graft-versus-leukemia (GvL)-effect^8^, demonstrating an unmet need for new treatment strategies for GvHD.

Recent research has highlighted correlations between the composition of the intestinal microbiome, GvHD pathophysiology, and clinical outcome after allo-SCT.^2,9,10^ The intestinal microbiome in allo-SCT recipients is altered, with reduced bacterial diversity and domination by single taxa, such as *Enterococcus.*^2,11–14^ Administration of broad-spectrum antibiotics and the conditioning therapy-derived toxicity are considered the main factors for profound microbiota disruptions during the peri-engraftment period.^6,15–17^ Another alteration that has emerged in recent studies is the depletion of microbial metabolites, which are produced as intermediates or end products of microbial metabolism.^18–22^ Those include short-chain fatty acids (SCFAs) and branched-chain fatty acids (BCFAs), which are derived from microbial fermentation of dietary compounds in the large intestine. Another class of metabolites are bile acids (BAs), which are synthesized in the liver as primary BAs and then metabolized or modified by the intestinal microbiota, forming secondary BAs. Metabolites within these two above mentioned classes are known to have immunomodulatory and tissue-homeostatic functions^23–25^ and to have protective effects on the incidence and severity of intestinal GvHD.^26^ For instance, a reduction of the SCFAs butyric and propionic acid during peri-engraftment is associated with a higher rate of GvHD in patients receiving allo-SCT.^26,27^ Previously, we established the *Immune-Modulatory Metabolite Risk Index (IMM-RI),* comprising butyric, propionic, and isovaleric acid, desaminotyrosine (DAT), and indole-3-carboxaldehyde (ICA). An *IMM-RI* low-risk, which is defined by high stool metabolite concentrations assessed at the peri-engraftment period, was associated with a better overall survival (OS) and a lower incidence of relapse in patients receiving alloSCT.^20^

Most studies have focused on microbiota composition and metabolite profiles during the peri-engraftment period, defined as the interval from day 0 to day +21–28, when hematologic reconstitution occurs.^28^ However, diversity and composition of the gut microbiome remain dynamic after engraftment.^11,27^ To date, only a few clinical studies have analyzed the microbiome and its metabolites beyond the peri-engraftment period, referred to as the “post-transplant period,” which usually ends at day +100, when the risk for cGvHD increases.^27,29^ Given the growing number of long-term allo-SCT survivors and their comorbidities,^30^ investigating persistent microbiome alterations is relevant, as a lasting intestinal dysbiosis has been associated with late complications such as cGvHD.^27,31,32^

We longitudinally tracked the intestinal microbiome via 16S rRNA gene amplicon sequencing and the microbial metabolome via mass spectrometry until day +100 in patients receiving alloSCT. We hypothesized that by the end of the post-transplant period, patients would show evidence of intestinal microbiome recovery. Accordingly, we expected that the restoration of bacterial populations would be accompanied by a parallel recovery of microbial metabolite profiles. Finally, we aimed to establish a microbiome signature that could serve as a biomarker for predicting clinical outcomes, such as OS, the onset of cGvHD, and relapse in the post-transplant period.

## METHODS

### Patient cohorts

In this two-centric, longitudinal, observational study, we included a *discovery cohort* (n = 20) from TUM and an independent *validation cohort* (n = 100) from UKR. The *discovery cohort* comprised patients who met the inclusion criteria of having at least one sample at each predefined time point: *PRE-TRANSPLANT* (*TX*, day −7), *PERI-ENGRAFTMENT* (day 0 to day +28), *EARLY POST-TX* (day +50 to day +92), and *LATE POST-TX* (day +93 or later, **Figure 1A**). The *validation cohort* included patients with at least one sample at *PRE-TX* and *LATE POSTTX* (**Figure 1B, Figure 1C**). All patients underwent allo-SCT for hematologic malignancies or myeloproliferative neoplasia and were treated per standard-of-care. The study was registered in the German Trial Register under DRKS00034175. Written informed consent was obtained from all participants in accordance with study protocols approved by the institutional review boards of TUM (295/18 S) and UKR (14-47_1-101, 14-101-0047, 21-2521-01). Patients were enrolled between 2019—2022 (TUM) and 2019—2023 (UKR). Acute GvHD was graded according to MAGIC criteria, with steroid-refractory disease defined as progression within 3 days of treatment initiation or lack of response within 5–7 days to prednisolone-equivalent doses ≥ 2 mg/kg/day.^29^ Mild GI-aGvHD comprised stages I–II, and severe GI-aGvHD stages III–IV. Chronic GvHD was graded according to NIH consensus criteria.^33^

**Figure 1:**
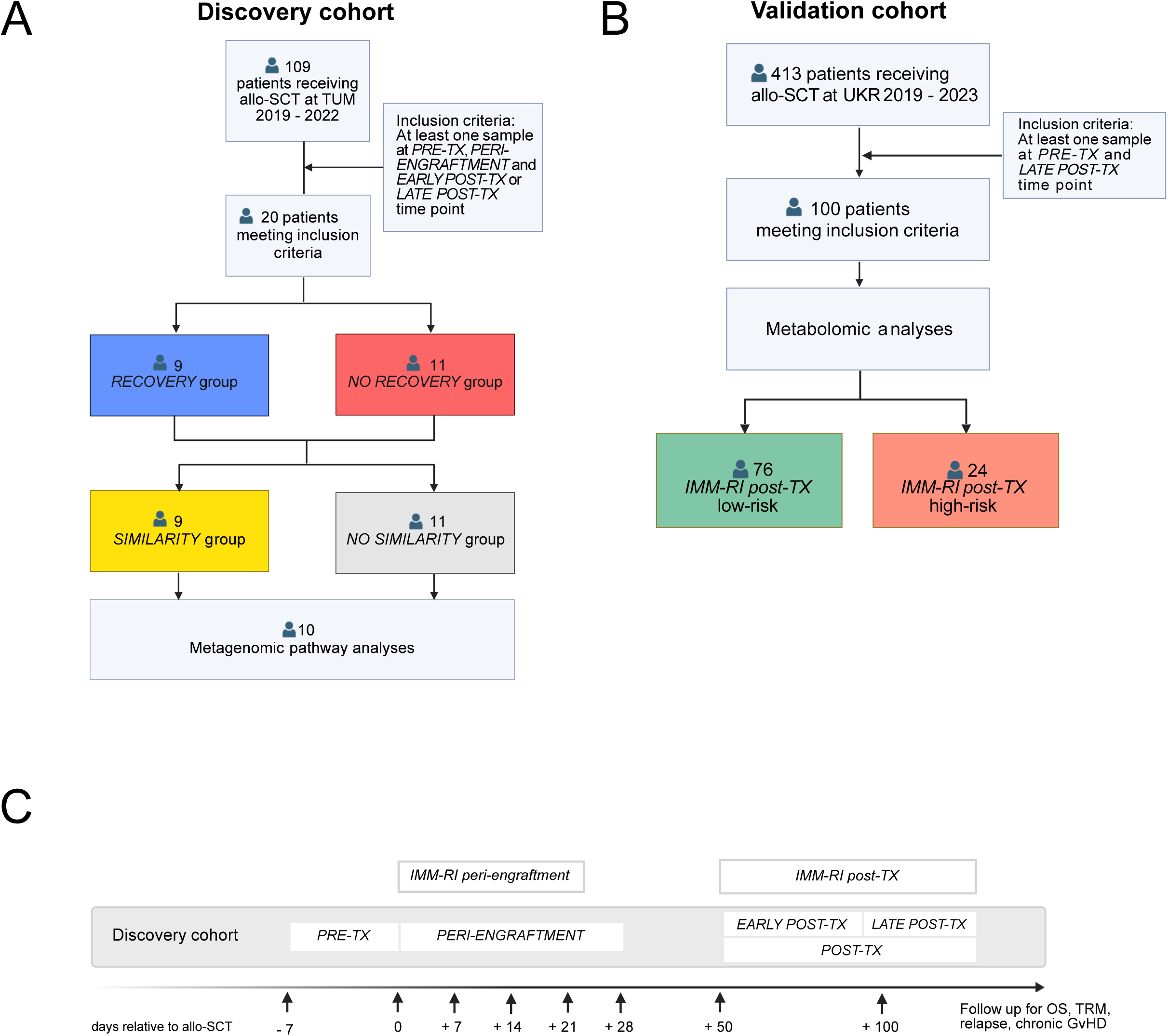
Consort charts, and sampling time points. **A:** *Discovery cohort*: Of 109 patients receiving allo-SCT at TUM between 2019-2022, 20 patients met the inclusion criteria of having at least one sample available at the *PRE-TX*, *PERIENGRAFTMENT*, *EARLY POST-TX,* and *LATE POST-TX* time points. Nine patients were stratified in the *RECOVERY* group and eleven patients in the *NO RECOVERY* group. In the second stratification, nine patients were stratified in the *SIMILARITY* group and eleven patients in the *NO SIMILARITY* group. Metagenomic pathway analyses were performed in a total of ten patients across both groups. Created with BioRender.com. **B:** *Validation cohort*: Of 413 patients receiving allo-SCT at UKR between 2019-2023, 100 patients met the inclusion criteria of having at least one sample available at the *PRE-TX* and *LATE POST-TX* time point. After stratification by *IMM-RI post-TX*, 76 patients were assigned to *IMM-RI post-TX* low-risk and 24 to *IMM-RI post-TX* high-risk. Created with BioRender.com. **C**: Overview of sampling time points and assessment of *IMM-RI peri-engraftment* and *IMM-RI post-TX: Discovery cohort*: Samples were collected at four time points *PRE-TRANSPLANT* (TX, day -7), *PERI-ENGRAFTMENT* (day 0 – day +28), *EARLY POST-TX* (day +50 – day +92) and *LATE POST-TX* (day +93 or later). *EARLY POST-TX* and *LATE POST-TX*, are described as one *POST-TX* time point to simplify presentation. *Validation cohort*: Samples were collected at two time points *PRE-TX* and *LATE-POST-TX*. The *IMM-RI peri-engraftment* was calculated using samples obtained between day +7 and day + 21 and was assessed only in the *discovery cohort*. The *IMM-RI post-TX* was derived from samples collected during *EARLY* and *LATE POST-TX* time points and was assessed in both cohorts. The time axis is schematic and not drawn to scale. Created with BioRender.com.

### Biospecimen collection and timepoints

Stool samples were collected at predefined calendar-driven time points from inpatients (day −7, 0, +7, +14, +21, +28) and outpatients (day +50, +100), within ±1–2 days of the scheduled date. Additional event-driven samples were obtained at the onset of GI-aGvHD. Detailed sampling and storage procedures are described in the **Extended Methods**.

Based on sampling time points, we defined four time points relative to allo-SCT to longitudinally assess taxonomic and metabolomic dynamics with a focus on post-transplantation microbiome recovery: *PRE-TRANSPLANT* (*TX*, day −7 to −1), *PERI-ENGRAFTMENT* (day 0 to +28), *EARLY POST-TX* (day +50 to +92), and *LATE POST-TX* (day +93 or later). These intervals reflect distinct clinical phases characterized by differences in immunosuppression, hematologic recovery, immune reconstitution, inor outpatient setting, and the transition from acute to chronic GvHD.

### Group stratification

Patients with an effective richness value, calculated as the mean of *EARLY* and *LATE POSTTX* values, that exceeded the 25^th^ percentile of the baseline value measured at *PRE-TX* were classified as part of the *RECOVERY* group. All other patients were assigned to the *NO RECOVERY* group (**Figure 2 B**). Of note, *RECOVERY* vs. *NO RECOVERY* refers to the intestinal microbiome and should not be confused with hematological recovery. Please refer to the discussion for a rationale.

**Figure 2:**
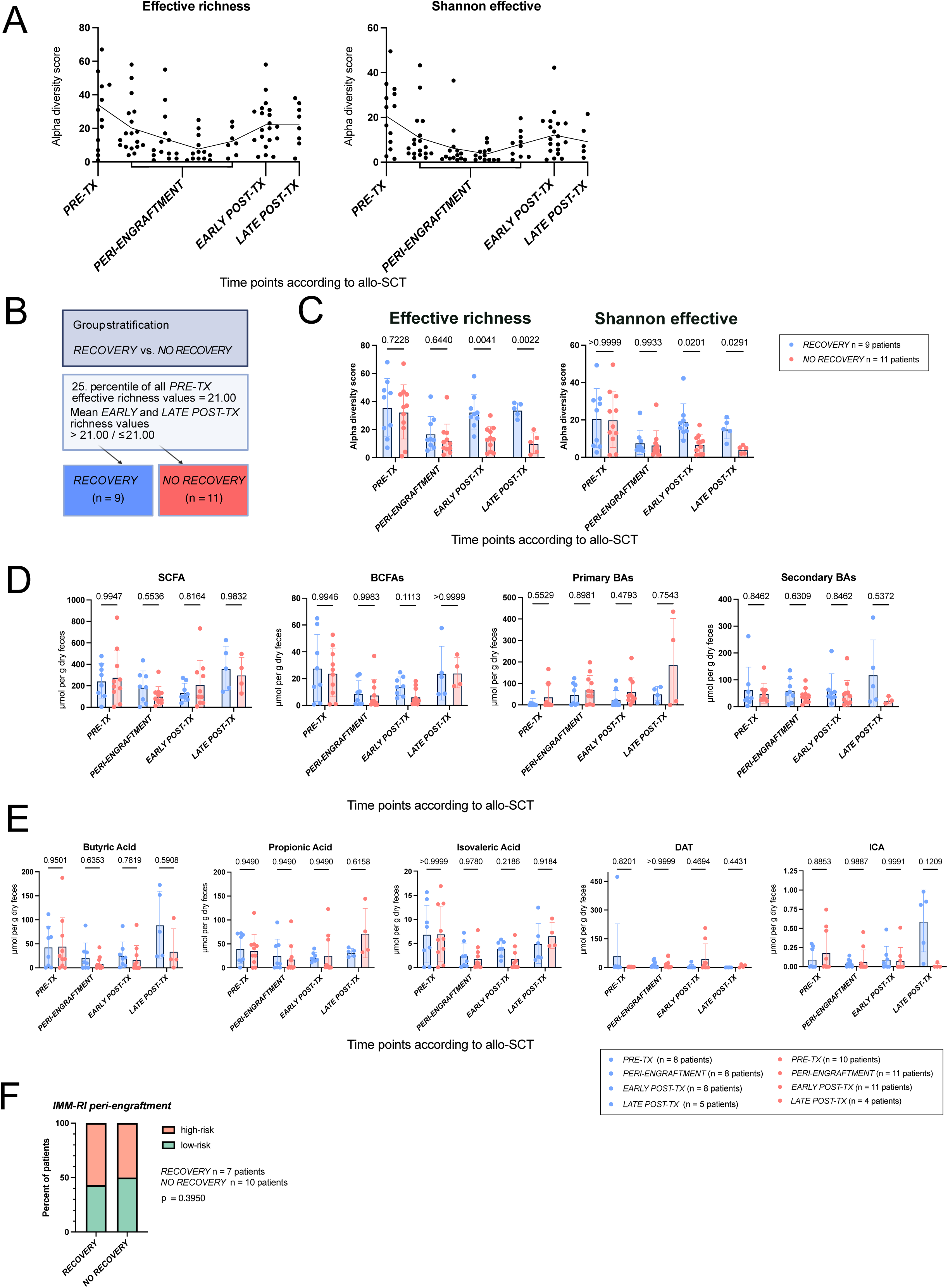
Stratification of patients receiving allo-SCT according to bacterial alpha diversity into *RECOVERY* vs. *NO RECOVERY* groups is not associated with a restoration of microbial metabolites during the *post-TX* period. **A**: Intestinal microbial composition at days relative to allo-SCT (x-axis) by alpha diversity (effective richness and Shannon effective numbers by observed Operational Taxonomic Units; yaxis). Each dot represents one individual patient sample. The line shows the means at specific time points. **B**: Illustration of group stratification based on the 25^th^ percentile of all *PRE-TX* effective richness values, i.e., 21.00. Patients were stratified into the *RECOVERY* group (n = 9), when their averaged effective richness value for the *EARLY* and *LATE POST-TX* was above 21.00, otherwise into the *NO RECOVERY* group (n = 11). Created with BioRender.com. **C**: Comparison of the time points between *RECOVERY* and *NO RECOVERY* for effective richness and Shannon effective numbers. The bars show mean values. Error bars indicate standard deviation. Points indicate individual patient samples. We applied repeated-measures ANOVA with the Geisser-Greenhouse correction and a mixed-effects analysis with multiple-comparison testing. Further, we corrected for multiple comparisons using Holm-Sidak. P-values < 0.05 were considered significant. **D**: Stool metabolite concentrations (in mol per gram of dried feces) according to time points relative to allo-SCT in *RECOVERY* (blue) and *NO RECOVERY* (red) groups for selected metabolite classes: SCFAs, BCFAs, primary BAs, and secondary BAs. The bars show the mean values. Error bars indicate standard deviation. Statistics as in **C**. **E**: Comparison of selected metabolites representing the *IMM-RI* As in D) according to time points in *RECOVERY* (blue) and *NO RECOVERY* (red) groups. The bars show the mean values. Error bars indicate standard deviation. Statistics are in **C**. **F**: Stratification of patients according to the *IMM-RI peri-engraftment* based on their recovery status (*RECOVERY vs*. *NO RECOVERY*). In *RECOVERY* there were three patients with *IMMRI peri-engraftment* low-risk (green) and four patients with *IMM-RI peri-engraftment* high-risk (red) values. *NO RECOVERY* included five patients with *IMM-RI peri-engraftment* high-risk (red) and five patients with *IMM-RI peri-engraftment* low-risk (green) values. 17 patients, of whom metabolite measurements were available, were included in the analysis. P-values < 0.05 were considered significant.

Patients were additionally stratified into *SIMILARITY and NO SIMILARITY* groups *as follows:* We calculated the beta diversity by percentage similarity of the mean of *EARLY and LATE POST-TX* values to the corresponding baseline of *PRE-TX* by using generalized UniFrac distance^34^ (gunif; **Extended Methods**). We used the mean of the similarity percentages (= 52.45%) as a cut-off for group stratification into a *SIMILARITY* (above the mean) and a *NO SIMILARITY* (below the mean) subgroup (**Figure 5 A**).

#### Immuno-modulatory metabolite risk index (IMM-RI)

The *IMM-RI* was assessed separately for two time points, the *peri-engraftment* and the *postTX* as previously established.^20^ Metabolite concentrations of five metabolites (propionic, isovaleric, and butyric acid, DAT, and ICA) were averaged from day +7 until +21 (*IMM-RI peri-engraftment*, as previously published^20^) and from day +50 until day +93 or later (*IMM-RI postTX)*. Previously published thresholds were applied for the *IMM-RI peri-engraftment*^20^ while for *IMM-RI post-TX*, optimized thresholds were determined using the Youden index (**Supplemental Table 1**). Concentrations below the respective cutoffs were assigned one point each, yielding an *IMM-RI* score of 0–5. Patients were classified as high-risk (3–5 points) or low-risk (0–2 points).

### 16S rRNA gene amplicon sequencing

Total DNA was isolated from stool samples and 16S rRNA gene amplicon sequencing (V3/V4 region) was performed using a two-step PCR library preparation followed by paired-end sequencing (MiSeq, Illumina). Raw reads were processed using the IMNGS2 pipeline based on UPARSE/UNOISE for quality control, chimera removal and zOTU clustering.^35–37^ Taxonomic assignment was performed using SILVA (v138.1) with taxonomy-informed clustering.^38^ Downstream analyses, including normalization and calculation of alpha and beta diversity metrics, were conducted in Rhea.^39^ Detailed laboratory protocols and bioinformatic procedures are provided in the **Extended Methods**.

### Metagenomics

Libraries were prepared from the same DNA extracts and sequenced on a NovaSeq platform (Illumina, PE150; target depth ∼5 Gbp/sample). Reads were quality filtered, trimmed and decontaminated prior to taxonomic profiling with MetaPhlAn v3.0^40^ and functional profiling using HUMAnN 3.^41^ A full description of library preparation, sequencing and computational analysis is provided in the **Extended Methods.**

### Metabolome analysis

Microbial metabolites, including SCFAs, BCFAs, tryptophan derivatives, desaminotyrosine, lactic acid, and primary and secondary BAs, were quantified in stool samples by targeted LC– MS/MS. Samples were extracted with internal standards and metabolite concentrations were normalized to stool dry weight. SCFAs and related metabolites were quantified using a 3-NPH derivatization method followed by UHPLC–MS/MS.^42^ BAs were measured using a targeted multiple-reaction-monitoring approach. Data processing included removal of features with high missingness, imputation at the limit of detection, and downstream statistical analysis in MetaboAnalyst.^43^ Comprehensive methodological details are provided in the **Extended Methods**. 16S rRNA gene sequencing, shotgun metagenomics, and targeted metabolomics data have been deposited in public repositories in accordance with journal guidelines. Accession numbers and reviewer access details are provided in the **Extended Methods** and will be publicly accessible upon publication.

### Statistical analyses

Microbiome alpha diversity (effective richness, Shannon effective) and beta diversity (generalized UniFrac (gunif))^44^ were calculated using the Rhea pipeline.^39^ Normality was assessed using standard distribution tests; as data were non-normally distributed, non-parametric Kruskal-Wallis tests followed by Dunn’s post-hoc tests were applied for group comparisons. For longitudinal metabolite data, generalized linear mixed-effects models (Gamma distribution, log link) were used, accounting for repeated measurements (random intercept) and interactions between disease severity and time points. Multiple testing was corrected via BenjaminiHochberg (FDR) or Holm-Šidák methods where appropriate. OS was estimated using the Kaplan-Meier method. Cumulative incidences of TRM, GvHD, and relapse were calculated using competing risk analysis.^45,46^ Statistical analyses were performed using R (lme4, emmeans, cmprsk packages) and GraphPad Prism. Significance was defined as p < 0.05. Detailed methodology, including specific formulas and normality testing procedures, is provided in the **Extended Methods**.

## RESULTS

### Patient characteristics of the *discovery cohort*

In this prospective, longitudinal, observational cohort of n = 20 patients undergoing allo-SCT at TUM, the mean age was 55 and 30% of the participants were females. Acute myeloid leukemia was the most common among a range of indications for allo-SCT. Most patients received reduced intensity conditioning (70%, n = 14) followed peripheral blood-mobilized stem cells (100%, n = 20) from a matched, unrelated donor (75%, n = 15). Over time, 75% (n = 15) of patients developed GI-aGvHD of any grade, many of which (67%, n = 10) suffered from steroid-refractory disease. No patients developed GI-cGvHD in the *discovery cohort* (**Table 1**). As epidemiological factors can alter microbial composition^47,48^ we assessed diet, alcohol consumption, body-mass index (BMI)^49^, physical activity, antibiotics, and immunosuppressive therapy^15,17^. All patients (n = 20) consumed a Western diet, 70% (n = 14) were smokers, 25% (n = 5) of patients consumed alcohol above the WHO-recommended levels, and half (n=10) were overweight or obese, while 75% (n = 15) were regularly physically active. There were no significant differences in epidemiological factors between the *RECOVERY* and the *NO RECOVERY* group (**Supplemental Table 2**). Antibiotic exposure was recorded as previously reported.^9^ *In PRE-TX* 20% (n = 3) antibiotics (BSA) before allo-SCT, and all (n = 20) during *PERIENGRAFTMENT* (**Supplemental Table 3**). Patients received GvHD prophylaxis according to institutional standards (**Supplemental Table 4**).

**Table 1:**
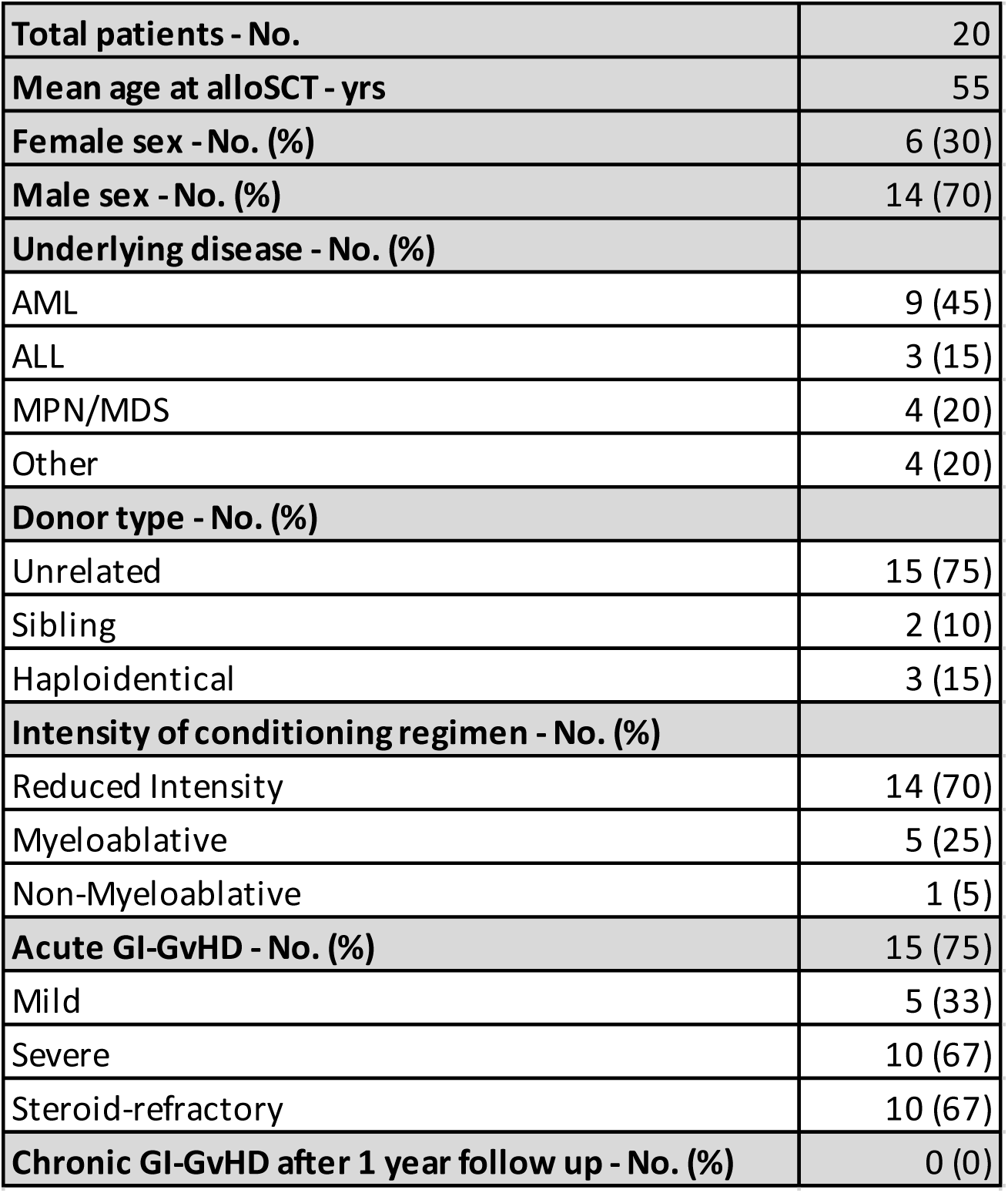
Patient characteristics: *Discovery cohort*. SCT, stem cell transplantation; AML, acute myeloid leukemia; ALL, acute lymphocytic leukemia; MDS, myelodysplastic syndrome; MPN, myeloproliferative neoplasia; GI-GvHD, gastrointestinal graft-versus-host disease. Diseases categorized as “other” include multiple myeloma, lymphoma, blastic plasmacytoid dendritic cell neoplasm, and T-prolymphocytic leukemia. According to MAGIC criteria, mild GvHD includes grades 0 to II; severe GvHD includes grades III to IV. Chronic GvHD is defined according to the National Institutes of Health 2014 consensus criteria.

|  |  |
| --- | --- |
| **Total patients - No.** | 20 |
| **Mean age at alloSCT - yrs** | 55 |
| **Female sex - No. (%)** | 6 (30) |
| **Male sex - No. (%)** | 14 (70) |
| **Underlying disease - No. (%)** |  |
| AML | 9 (45) |
| ALL | 3 (15) |
| MPN/MDS | 4 (20) |
| Other | 4 (20) |
| **Donor type - No. (%)** |  |
| Unrelated | 15 (75) |
| Sibling | 2 (10) |
| Haploidentical | 3 (15) |
| **Intensity of conditioning regimen - No. (%)** |  |
| Reduced Intensity | 14 (70) |
| Myeloablative | 5 (25) |
| Non-Myeloablative | 1 (5) |
| **Acute GI-GvHD - No. (%)** | 15 (75) |
| Mild | 5 (33) |
| Severe | 10 (67) |
| Steroid-refractory | 10 (67) |
| **Chronic GI-GvHD after 1 year follow up - No. (%)** | 0 (0) |

### Longitudinal dynamics of bacterial diversity in patients receiving allo-SCT

To analyze longitudinal changes in microbial composition, we evaluated alpha diversity using effective richness and Shannon effective. An overview of the sampling time points in both cohorts is provided in **Figure 1C**. Alpha diversity decreased from *PRE-TX* to *PERI-ENGRAFTMENT* with a nadir on day +14, then increased by *EARLY and LATE POST-TX* (**Figure 2A**).

Given the known association between peri-engraftment alpha diversity and outcomes in alloSCT^2^, we investigated whether recovery of the microbiome during the post-transplantation period may serve as a biomarker of long-term clinical outcomes. Therefore, we stratified patients based on microbial recovery between baseline (*PRE-TX) and EARLY and LATE POST-TX* into *RECOVERY* (n = 9, 45%) and *NO RECOVERY* (n = 11, 55%; **Figure 2B**) as detailed in the *Methods*. Consequently, alpha diversity in the *POST-TX* time points was significantly different between the *RECOVERY* and *NO RECOVERY* groups, but no difference was seen in *PRE-TX nor in PERI-ENGRAFMENT* (**Figure 2C**). As results of *EARLY* and *LATE POST-TX* were comparable, we henceforth describe both time points as *POST-TX* to simplify presentation.

### No differences in metabolite profiles between *RECOVERY* and *NO RECOVERY* groups

We next compared microbial metabolic output, including stool SCFAs, BCFAs, and primary and secondary BAs, between *RECOVERY* and *NO RECOVERY* groups (**Figure 2D, Supplemental Table 5**). Regardless of time point, there were no significant differences in stool concentrations of any metabolite class between groups.

We then analyzed only *IMM-RI* metabolites (butyric, propionic, isovaleric acid, DAT, ICA) and again observed no difference between *RECOVERY* and *NO RECOVERY* groups. Using linear regression analyses, we observed no significant correlations between stool concentrations of IMM-RI metabolites and effective richness values measured at *POST*-*TX* (**Figure 2E, Supplemental Figure 1A**).

Finally, the *IMM-RI peri-engraftment* was calculated for each patient.^20^ In the *RECOVERY* group, three patients were low-risk and four high-risk according to the *IMM-RI peri-engraftment* (i.e., high vs. low stool metabolite concentrations, respectively), whereas in *the NO RECOVERY* group, five patients were low-risk and five high-risk. No significant difference was observed between groups by Fisher’s exact test (**Figure 2F**). Overall, microbial recovery based on alpha diversity was not associated with restoration of *POST*-*TX* metabolite profiles.

### Bacterial ‘Recovery’ is a poor predictor of clinical outcomes

We next assessed whether bacterial *RECOVERY* status was associated with clinical outcomes. OS did not differ between groups (log-rank test; **Figure 3A**). *RECOVERY* was not significantly associated with relapse or TRM in the competing risk analysis (**Figure 3B**). In contrast, *NO RECOVERY* was independently associated with severe GI-aGvHD after accounting for the competing risk of death (**Figure 3C**). Thus, alpha diversity differences observed according to *RECOVERY* status were not generally reflected in clinical outcomes, except for GIaGvHD.

**Figure 3:**
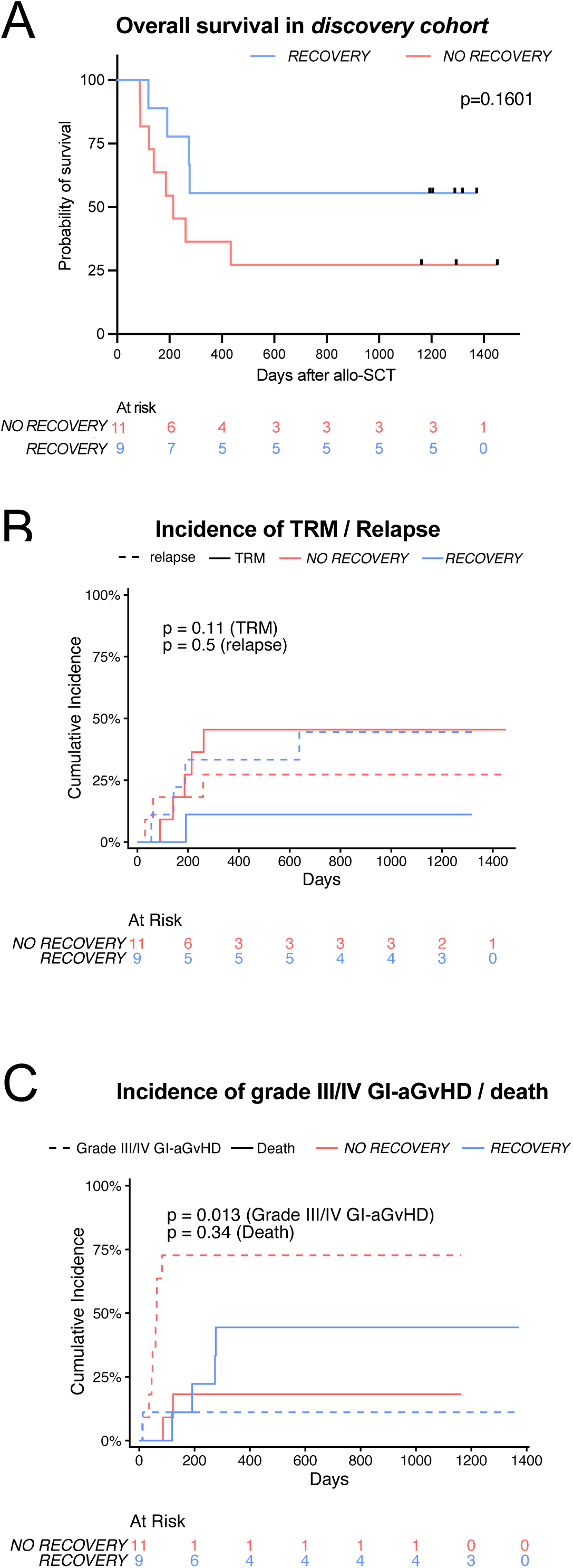
Clinical outcomes in allo-SCT patients stratified into *RECOVERY* vs. *NO RECOVERY groups*. **A**: OS according to *RECOVERY* (blue) *vs. NO RECOVERY* (red). Number at risk are indicated below: In the *RECOVERY* group, there were four deaths among nine patients, and for the *NO RECOVERY* there were eight deaths among eleven patients. Shown is the analysis via the Kaplan–Meier estimator and significance was assessed using the log-rank test. **B:** Cumulative incidence of TRM (solid line) and relapse (dashed line) as competing risks between patients stratified into *RECOVERY* (blue) and *NO RECOVERY* (red). Numbers at risk are in the table below. Cumulative incidence functions between groups were tested for equality using Gray’s test. P-values < 0.05 were considered significant. **C:** Cumulative incidence of grade III/IV GI-aGvHD (solid line) with death (dashed line) as competing risks between *RECOVERY* (blue) and *NO RECOVERY* (red). Numbers at risk are in the table below. Cumulative incidence functions between groups were tested for equality using Gray’s test. P-values < 0.05 were considered significant.

### Taxonomic composition and similarity between *PRE-TX* and *POST-TX* in individual patients

Alpha diversity metrics do not capture taxonomic composition. Given the limited association between *RECOVERY* status, microbial metabolite concentrations, and clinical outcomes, we hypothesized that microbiota associated with the production of immunomodulatory metabolites, present before transplantation, may have been lost after allo-SCT. To address this, we analyzed genus-level taxonomic profiles of all available stool samples (**Figure 4**) and described the community composition longitudinally in individual patients with beta diversity. To show changes in microbial composition between *PRE-TX* and *POST-TX,* we defined a *SIMILARITY* measure based on gunifbeta diversity as described above (**Figure 5A**; **Supplemental Table 6**). In the *RECOVERY* group, six patients (67%) demonstrated compositional similarity to baseline, whereas three (33%) did not (**Figure 4** left panel, **Figure 5B**). In the *NO RECOVERY* group, four patients (36%) retained baseline similarity, including two who showed transient loss of diversity during *PERI-ENGRAFTMENT* followed by compositional restoration (Patient M and T, **Figure 4**, right panel). The remaining seven patients (64%) exhibited persistent alterations in *POST-TX* community structure compared to baseline. Overall, recovery of bacterial richness, measured by alpha diversity, did not necessarily correspond to the restoration of baseline microbial composition, as assessed by beta diversity in the long term, which may explain the weak correlation between richness and metabolite concentrations (**Figure 2D, E**).

**Figure 4:**
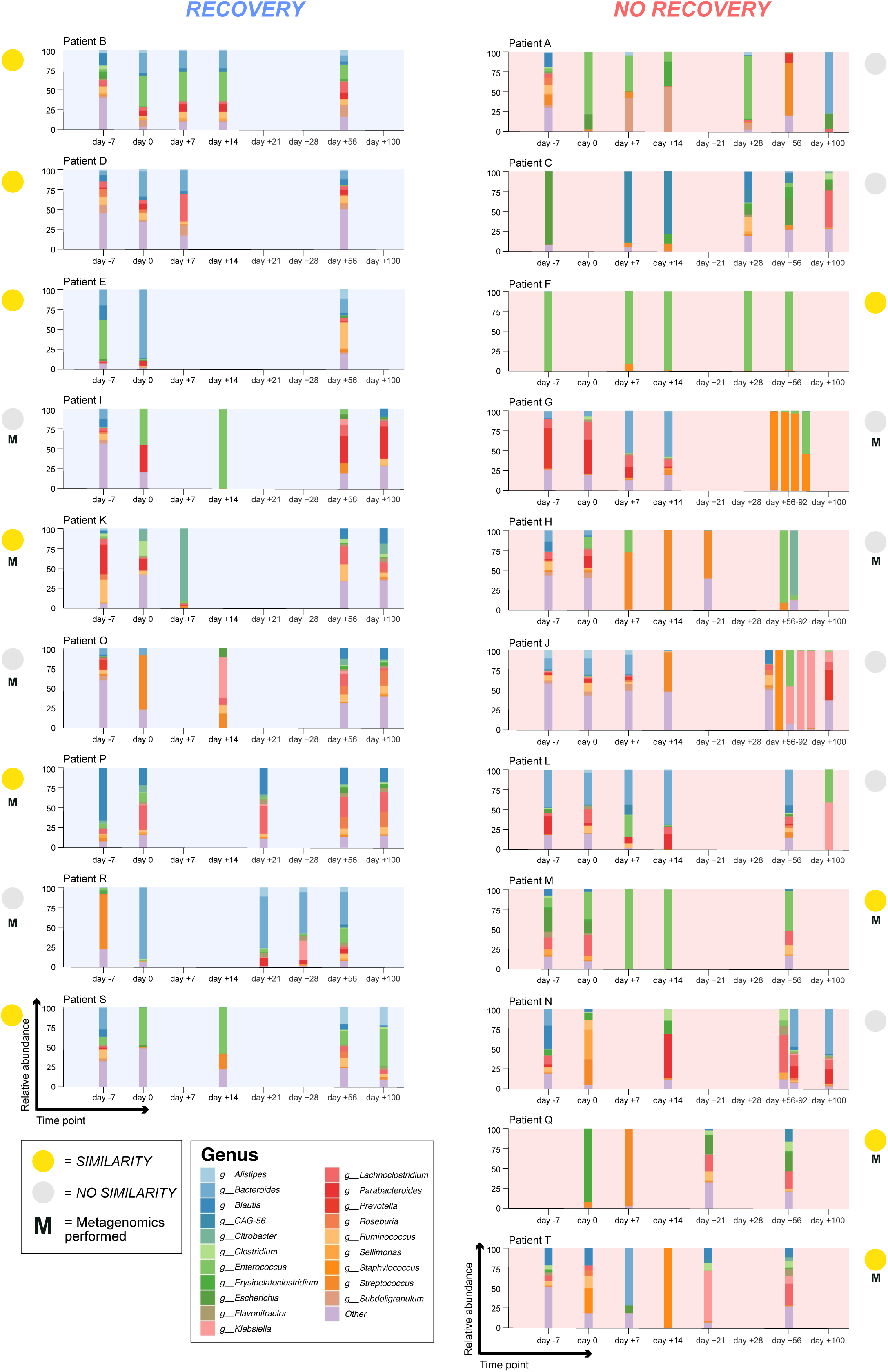
Longitudinal taxonomic compositions in individual patients according to *RECOVERY* vs *NO RECOVERY*. Illustration of taxonomic compositions in individual patient samples stratified into *RECOVERY* (n = 9, blue) and *NO RECOVERY* (n = 11, red). Relative abundance of bacterial genera is indicated on the y-axis, time points relative to allo-SCT from day -7 to day +100 on the x-axis. Taxonomic compositions are shown at genus level. A legend of the 20 most abundant genera is given. Circles on each diagram show group affiliation, i.e., S*IMILARITY* (yellow) and *NO SIMILARITY* (gray). “M” (black) indicates that metagenomics was performed.

**Figure 5:**
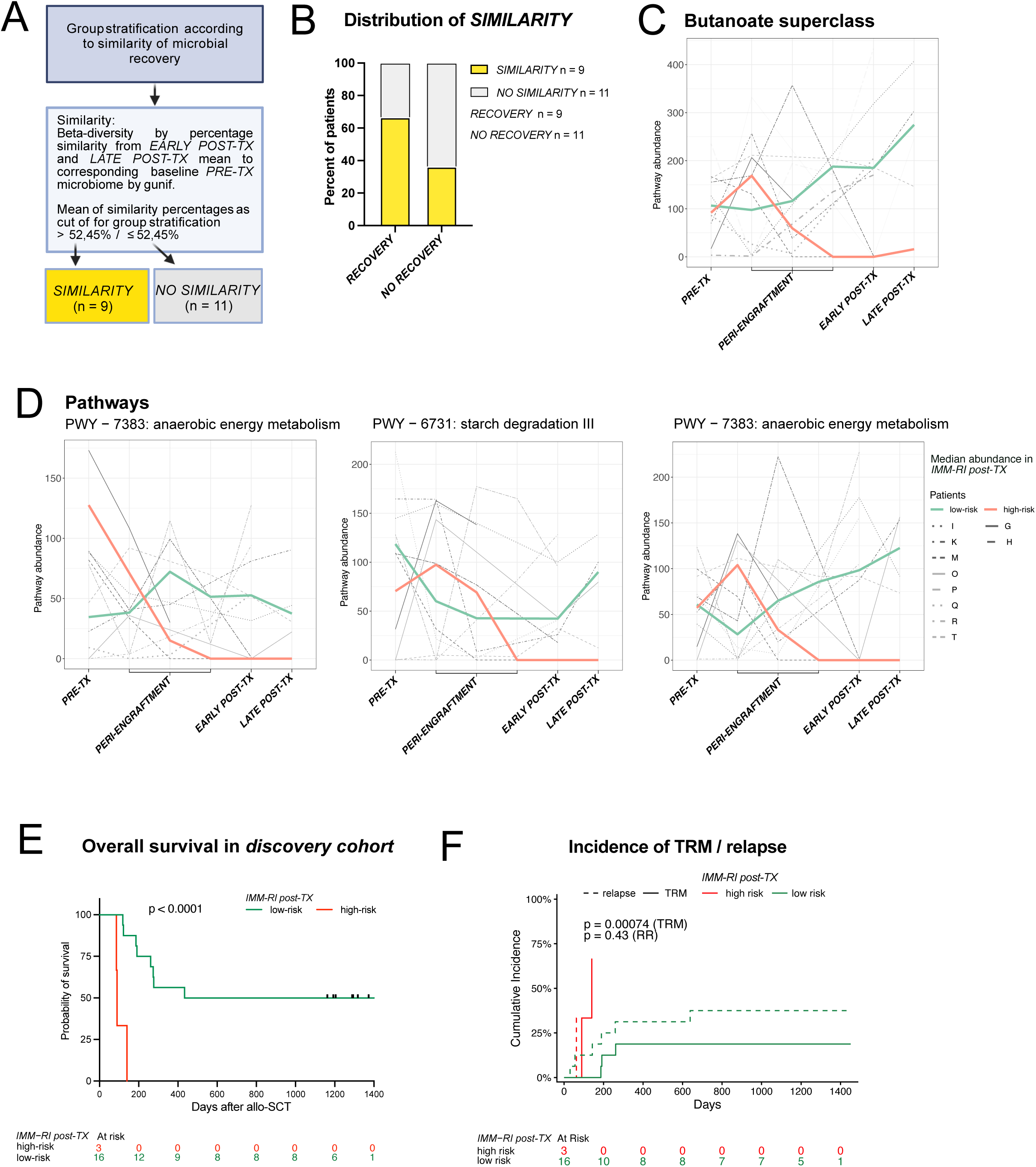
S*I*MILARITY *according to RECOVERY status and* metagenomic profiles, and clinical outcomes in patients *stratified according to IMM-RI post-TX (discovery cohort).* **A:** Illustration of group stratification based on each individual’s *SIMILARITY* of the mean microbial composition at *POST-TX* to the corresponding baseline microbiome (*PRE-TX)* using the beta diversity measure gunif. The mean of the similarity percentages was used as a cutoff for group stratification into *SIMILARITY* (n = 9, yellow) and *NO SIMILARITY* (n = 11, gray). Created with BioRender.com. **B**: In *RECOVERY* six (67%) patients were stratified into *SIMILARITY* (yellow) and three (33%) patients into *NO SIMILARITY* (gray). In *NO RECOVERY* group, four (36%) patients were stratified into *SIMILARITY* (yellow) and seven (64%) patients into *NO SIMILARITY* (gray). **C D**: Longitudinal dynamics of the abundance butyric-acid superclass pathways (**C**), relevant pathways for butyric acid synthesis (**D**) assessed via metagenomic sequencing, for eight patients from *IMM-RI post-TX* low-risk (I, K, M, O, P, Q, R, and T) and two patients from *IMM-RI post-TX* high-risk (G, and H). The green and red line show the group medians. Abundance is indicated on the y-axis and time points relative to allo-SCT from day -7 until day +100 on the x-axis. **E**: OS between *IMM-RI post-TX* high-risk and *IMM-RI post-TX* low-risk patients in the discovery cohort. *IMM-RI post-TX* was calculated from mean of metabolite concentrations from day +50 until day +100. Analysis was performed with the Kaplan–Meier estimator; significance was determined by the log-rank test. Numbers at risk are indicated in the table below. P-values < 0.05 were considered significant. 19 patients, of whom metabolite measurements were available, were included in the analysis. **F:** Cumulative incidence of TRM (solid line) with relapse (dashed line) as a competing risk between the *IMM-RI post-TX* low-risk (green) and *IMM-RI post-TX* high-risk (red) group. There were two deaths attributed to TRM and one death attributed to relapse among three patients in the *IMM-RI post-TX* high-risk group. Numbers at risk are indicated in the table below. Pvalues < 0.05 were considered significant.

### *IMM-RI post-TX low-risk* is associated with an increased abundance of butyric acid synthesis pathways and better OS

Given that 33% of patients in the *RECOVERY* group returned to their baseline microbiome community composition, we hypothesized that microbial metabolic readouts, such as biosynthetic pathways or metabolite concentrations at *POST-TX*, might better predict clinical outcomes. We therefore adapted the *IMM-RI* from the peri-engraftment to the *POST-TX* time point (see *Methods*), stratified patients into *IMM-RI post-TX* lowand high-risk groups and performed longitudinal stool metagenomic sequencing in n=10 patients. We focused on superclass pathways and butyrate-associated pathways, given the established role of butyric acid in immune modulation, T_reg_ expansion, microbiome homeostasis after allo-SCT, ^24,25,50^ and aGvHD prevention and modulation.^18,23,51,52^ Consistent with our prior findings in the peri-engraftment setting,^20^ patients with *IMM-RI post-TX* low-risk showed higher abundance of superclass pathways (**Figure 5C, Supplemental Table 7**) and of PWY-7383 (anaerobic energy metabolism), PWY-6731 (starch degradation III), and PWY-5676 (acetyl-CoA fermentation to butanoate II; **Figure 5D**), suggesting preserved butyrate biosynthesis capacity.

We next assessed the association with clinical outcome: *IMM-RI post-TX* was significantly associated with OS (**Figure 5E**), with low-risk patients showing improved OS compared with high-risk (HR 0.1156, 95% CI 0.005981 – 2.234, p < 0.0001). TRM was significantly lower in *IMM-RI post-TX* low-risk patients after adjustment for the competing risk of relapse (**Figure 5F**), whereas the incidence of severe GI-aGvHD did not differ between groups (**Supplemental Figure 2A**).

### *Validation cohort*: the *IMM-RI post-TX* associates with OS, relapse and cGvHD incidence

To validate our findings, we tested the *IMM-RI post-TX* in an independent *validation cohort* of n = 100 patients receiving allo-SCT at UKR (**Figure 1B**). Patient characteristics are summarized in **Table 2**. The mean age at allo-SCT was 56 years and 39% of the participants were females. Acute leukemia represented the most common indication for allo-SCT. Most patients received reduced-intensity conditioning (86%) and underwent transplantation from a matched unrelated donor (75%). Overall, 60% of patients developed aGvHD, of whom 60% presented with GI-aGvHD. In total, 66% developed cGvHD,^33^ with 61% occurring after prior aGvHD and 39% arising *de novo*. Antibiotic exposure during *PRE-TX* and *PERI-ENGRAFTMENT* differed significantly between *IMM-RI post-TX* lowand high-risk (Fisher’s exact test, p = 0.019), primarily due to a lower proportion of antibiotic-free patients in the high-risk group (**Table 3**).

**Table 2:** Patient characteristics: *Validation cohort*. Descriptions, see Table 1. Diseases categorized as “other” include lymphoma and severe aplastic anemia.

|  |  |
| --- | --- |
| **Total patients - no.** | 100 |
| **Mean age at alloSCT - yrs.** | 56 |
| **Female sex - no.** | 39 |
| **Male sex - no.** | 61 |
| **Underlying disease - no.** |  |
| Acute leukemia | 52 |
| MDS/MPN | 30 |
| Other | 18 |
| **Donor type - no.** |  |
| Unrelated | 75 |
| Sibling | 10 |
| Haploidentical | 15 |
| **Intensity of conditioning regimen - no.** |  |
| Reduced Intensity | 86 |
| Myeloablative | 13 |
| Non-Myeloablative | 1 |
| **Acute GvHD\*\* no.** | **60** |
| Acute GI-GvHD no. (%) | 36 (60) |
| **Chronic GvHD\*\* no.** | **66** |
| Acute and chronic GvHD no. (%) | 40 (61) |
| Only chronic GvHD no. (%) | 26 (39) |
| Chronic Skin GvHD no. (%) | 48 (73) |
| ** any organ or severity considered |  |

**Table 3:** Beginning of antibiotic treatment according to *IMM-RI post-TX* low-risk and high-risk in *validation cohort*. Distribution of broad-spectrum antibiotic (BSA) administration, including piperacillin-tazobactam and carbapenems, by timing (*PRE-TX* and *PERI-ENGRAFTMENT*) and by *IMM-RI postTX* lowand high-risk. BSA administration that started in the *PRE-TX* time point and continued into *PERI-ENGRAFTMENT* was counted only in *PRE-TX*. The overall distribution of BSA use differed significantly between *IMM-RI post-TX* low-risk and high-risk patients (Fisher’s exact test, p = 0.019).

| No. patients (%) | *IMM-Rl post-TX* |  |
| --- | --- | --- |
|  | low risk | high risk |
| No BSA | 12 (16) | 1 (4) |
| BSA in *PRE-TX* | 11 (14) | 4 (17) |
| BSA in *PERI-ENGRAFTMENT* | 53 (70) | 19 (79) |
| Total | 76 (100) | 24 (100) |
| p-Value | 0.019 |  |

Kaplan–Meier analysis showed that *IMM-RI post-TX* low-risk status was associated with improved OS (HR 0.2052, 95% CI 0.07703 – 0.5466, p < 0.0001), consistent with the discovery cohort (**Figure 6A**). The *IMM-RI post-TX* predicted OS after two years with an area under the curve of 0.731 (95% CI: 0.616−0.846), a negative predictive value of 0.881 and a positive predictive value of 0.542 (**Supplemental Figure 3B**). *IMM-RI post-TX* was also significantly associated with relapse-free survival (**Figure 6B**). A higher relapse-related mortality with increased cumulative incidence was observed in patients with *IMM-RI post-TX* high-risk when accounting for TRM as a competing risk (**Figure 6C**).

**Figure 6:**
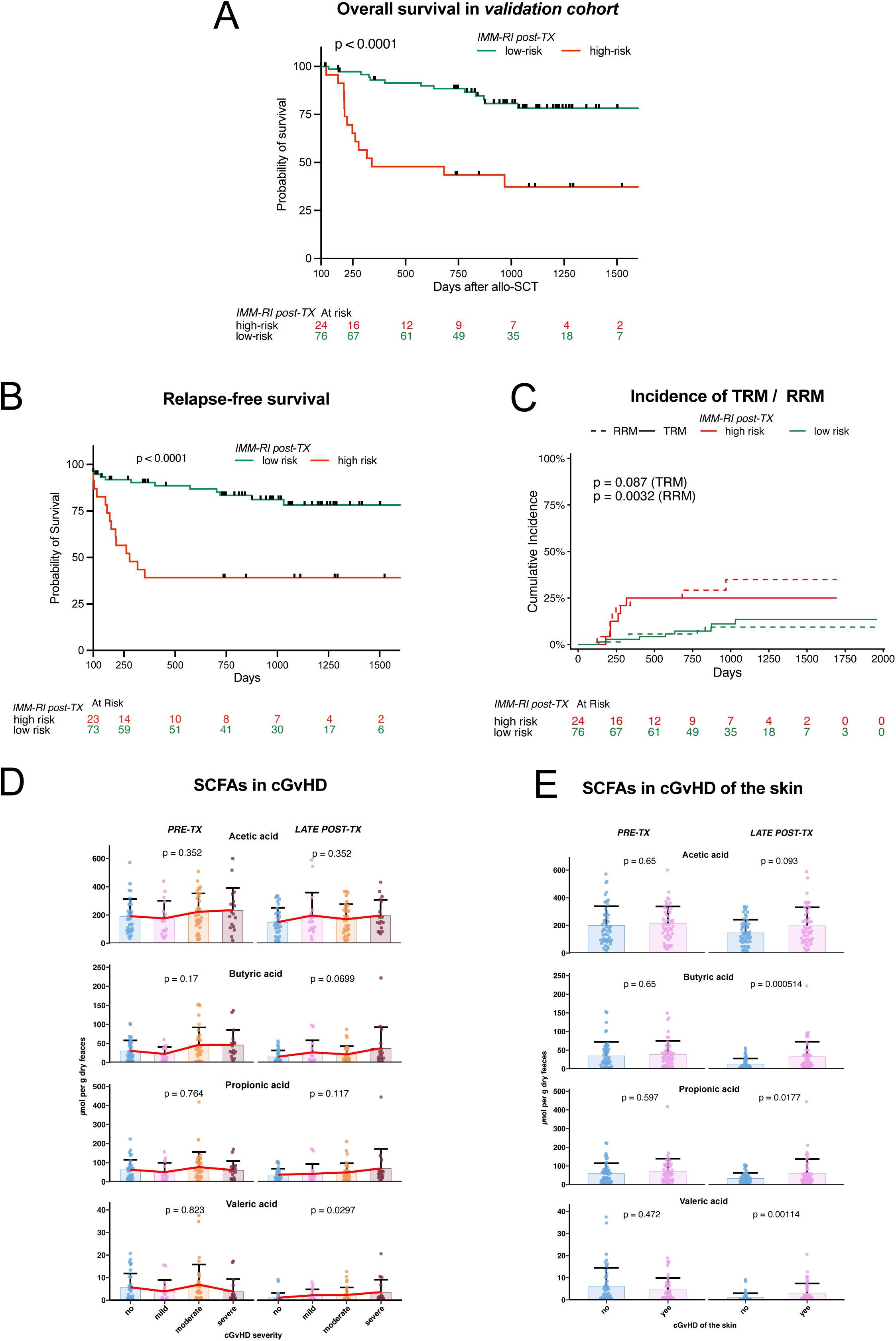
IMM-RI post-TX predicts clinical outcomes in the validation cohort, and SCFA concentrations are increased in cGvHD. **A:** OS between *IMM-RI post-TX* high-risk (n = 24) and *IMM-RI post-TX* low-risk (n = 76) patients. OS is shown from day +100 onward, reflecting the inclusion criteria of survival at *LATE POST-TX*. There were 13 deaths in the *IMM-RI post-TX* low-risk and 14 deaths in *IMM-RI postTX* high-risk, respectively. Survival analysis was performed using the Kaplan–Meier method and statistical significance was assessed by the log-rank test. Numbers at risk are indicated in the table below. P values < 0.05 were considered statistically significant. **B**: Relapse-free survival between *IMM-RI post-TX* high-risk (n = 24) and *IMM-RI post-TX* lowrisk (n = 76) patients. Relapse-free survival is shown from day +100 onward, reflecting the inclusion criteria of survival at *LATE POST-TX*. Statistics are in **A**. **C**: Cumulative incidence of TRM (solid line) with relapse-related mortality (RRM) (dashed line) as a competing risk between the *IMM-RI post-TX* low-risk (green) and *IMM-RI post-TX* highrisk (red) group. TRM and RRM take into account a follow-up period of one year. Cumulative incidence functions between groups were tested for equality using Gray’s test. P-values < 0.05 were considered significant. **D:** Stool concentration of SCFAs (in µmol per gram of dried feces) acetic, butyric, propionic, and valeric acid (VA) according to cGvHD grading (no, mild, moderate, severe in blue, pink, yellow, brown, respectively) and time point (*PRE-TX* and *LATE POST-TX*). Statistical significance was assessed to determine whether metabolite concentrations increased with increasing GvHD severity. The bars show the mean values. Red lines connect mean values across severity groups. Error bars indicate standard deviation. Significance was assessed using generalized linear mixed-effects models (Gamma family, log link) with a random intercept for subject; effects at each timepoint were estimated and tested. P-values were corrected for multiple testing using Benjamini-Hochberg false discovery rate (FDR). Points indicate individual patient stool samples sampled at the specified time points. P-values < 0.05 were considered significant. **E:** SCFAs acetic, butyric, propionic, and valeric acid according to incidence of cGvHD of the skin (yes vs. no in pink vs. blue, respectively) and time point (*PRE-TX* and *LATE POST-TX*). Statistics are in **D**.

Given the well-established concept that an enhanced GvL effect, while reducing relapse rates, may increase the risk of cGvHD and based on the emerging evidence of context-dependent stimulatory vs. suppressive effects of metabolites, specifically SCFAs,^53^ we analyzed metabolite concentrations according to cGvHD organ manifestation and severity. Generalized linear mixed-effects modeling revealed no association between SCFA levels and cGvHD severity at *PRE-TX*. However, at *LATE POST-TX*, valeric acid (VA) concentrations increased significantly with increasing cGvHD severity (**Figure 6D**). As the skin is the most frequently affected organ in cGvHD, we further analyzed this subgroup and observed significantly elevated levels of VA, butyric, and propionic acid in patients with skin cGvHD at *LATE post-TX* (**Figure 6E**). Collectively, these findings validate the *IMM-RI post-TX* as a prognostic marker associated with survival and relapse related mortality and highlight the potential role of microbially-derived IMMs in post-transplant immune modulation.

## DISCUSSION

In this study, we analyzed taxonomic shifts in the intestinal bacteriome and the dynamics of metabolite profiles in the late post-transplant period. We found a decline in bacterial richness values during *PERI-ENGRAFTMENT,* consistent with findings from national^20^ and international cohorts^2^ (**Figure 2A**). While several studies have assessed microbial diversity shortly after allo-SCT, few have examined later time points, leaving the longitudinal dynamics of bacterial recovery largely unexplored. Riwes et al. reported an increase in Shannon diversity at day +100 after allo-SCT, without comparison to baseline.^52^ Similarly, Bansal et al. showed that alpha diversity returns to baseline by day +100, albeit with substantial inter-patient variability.^17^ Patients experiencing aGvHD show a greater diversity loss at day +130 compared to other patients.^32^ Rashidi et al. observed a slight increase in Shannon diversity over nine months post-transplant, but levels never returned to baseline. ^54^ Other studies report persistent loss of diversity for up to 12 months or even 10 years after allo-SCT. ^27,31^ Notably, most of these studies lack a reference value to compare post-transplant microbial structure to baseline. Ingham et al. assessed microbial recovery at three months relative to pre-transplant levels, but without defining “recovery.”^55^ To address this, we established a permissive cutoff by stratifying patients at the 25^th^ percentile of bacterial recovery into *RECOVERY* and *NO RECOVERY* groups (**Figure 2B**) and tested associations with metabolite concentrations and clinical outcomes. Surprisingly, metabolite concentrations did not differ between groups (**Figure 2D, E**), and bacterial recovery was not associated with OS or TRM (**Figure 3A, B**), consistent with Shtossel et al., who found post-transplant microbial composition to be a less reliable predictor of clinical outcomes than peri-engraftment. ^56^ However, *NO RECOVERY* was significantly associated with severe GI-aGvHD (**Figure 3C**), and several patients exhibited dominance by single taxa a known feature of dysbiosis during GI-aGvHD. ^11,57^

We observed that bacterial recovery does not imply a return to the baseline microbial composition (**Figure 4**). This finding is consistent with observations in a pediatric cohort, in which the majority of patients did not revert to their initial microbial profile.^58^ The inability to maintain specific taxa that may be important for metabolite production could explain the weak correlation between bacterial and metabolite recovery. However, it remains debatable whether patients should be expected to return to their *PRE-TX* composition, as many may already exhibit an altered microbiome due to prior exposure to broad-spectrum antibiotics, repeated cycles of chemotherapy, and potentially refractory disease.^59^ Alternatively, the microbiome may stabilize into a new, community structure rather than reverting to its original state.

Analyzing alpha diversity alone does not provide sufficient context about community composition or metabolic capacity, and biomarkers based on taxonomy-based microbiome signatures have proven difficult to reproduce across cohorts due to the heterogeneous and highly individual nature of the microbiome.^60^ Therefore, we shifted our focus to microbial metabolites. Given that the *IMM-RI peri-engraftment* correlates with clinical outcomes after allo-SCT^20^, and since evidence suggests that metagenomics and metabolomics in the peri-engraftment better predict clinical outcomes^61^, we hypothesized that metabolite profiles may more effectively predict clinical outcomes at later time points. To test this, we adapted the *IMM-RI* to the *post-TX* period (**Figure 5C, D**) and observed that the *IMM-RI post-TX* correlates strongly with OS (**Figure 5E**) and reduced TRM (**Figure 5F**). We focused on butyric acid synthesis, as butyric acid plays a critical role in immune regulation by inducing T_regs_ in the colon and suppressing inflammation.^62^ We recognized that patients with *IMM-RI post-TX* low-risk were able to maintain abundance of butyric acid synthesis pathways during PERI-ENGRAFMENT and POST-TX timepoints (**Figure 5C, D**). This is consistent with preservation of butyrate-producing bacteria during the peri-engraftment period, which has been linked to improved OS^51^ and a reduced incidence of both acute and cGvHD.^18,26,27^

The clinical and biological relevance of microbiota-derived metabolites at day +100 remains poorly defined, particularly because existing studies have reported heterogeneous findings and because serum or plasma metabolite measurements may not adequately reflect the intestinal metabolite milieu captured in stool: One study reported that SCFA-producing bacteria, which are considered protective against GvHD^51^, were reduced by day +100.^17^ Another study demonstrated protective effects of serum butyric and propionic acid concentrations on day +100 for the development of cGvHD^27^. However, SCFAs measured in serum or plasma differ from those detected in fecal samples. For example, butyric acid is largely consumed locally by colonocytes and only minimally absorbed into the circulation.^63^ Other SCFAs, such as propionate, are absorbed but rapidly metabolized in the liver, resulting in low blood concentrations.^50^ In other hematologic malignancies, higher stool butyric acid concentrations and the maintenance of butyrate-producing bacteria by day +100, after autologous stem cell transplantation or induction therapy, have been associated with sustained minimal residual disease negativity.^64,65^ Beyond the immunomodulatory role of SCFAs^66^ and their potential to reduce GvHD,^18,23,27^ other metabolites such as DAT, a component of the *IMM-RI*, affect clinical outcomes. Peri-engraftment stool DAT concentrations correlate with improved OS and reduced relapse rates in patients receiving allo-SCT.^67^ Experimentally, DAT protects intestinal epithelial cells from acute damage, improves GvHD survival, and promotes effector T cells, enhancing the GvL-effect.

To ensure generalizability, findings from the *discovery cohort* were validated in an independent cohort (**Figure 1 B**), confirming the significance of *IMM-RI post-TX* (**Figure 6 A**). The absence of an association between TRM and *IMM-RI post-TX* (**Figure 6 C**) is most likely attributable to selection bias, as only patients surviving beyond day +100 were included in the analysis. Patients with *IMM-RI post-TX* low-risk exhibited a longer relapse-free survival and a lower relapse related mortality (**Figure 6B,C**) supporting the idea of an immunostimulatory role of microbial metabolites in enhancing the GvL effect.

Conditioning and allo-SCT induced intestinal tissue and microbiome -damage drive dysbiosis, which is further aggravated by antibiotic exposure. This microbial disruption promotes a proinflammatory milieu, impairs T_reg_ differentiation disrupts immune tolerance.^68,69^ A loss of tolerogenic signals favors the activation of alloreactive donor T cells and the onset of GvHD.^3,8,70^ We propose a context-dependent, dual role for microbial metabolites: During the peri-engraftment period, specific SCFAs may exert protective effects on the intestinal barrier, mitigating aGvHD.^18,23,26,51^ In contrast, in the late post-transplant period, elevated SCFAs may support effector T cell function, thereby enhancing GvL and reducing relapse, albeit at the cost of increased risk of cGHVD. This model may also help explain findings from the French ALLOZITHRO cohort, in which prolonged azithromycin exposure after allo-SCT was associated with impaired T-cell function, and an increased risk of relapse. ^71^ One possible interpretation is that antibiotic-mediated depletion of microbiota-derived metabolites may further dampen antitumor immunity by limiting the metabolic support required for effective T-cell effector function. In a prospective study, prebiotic supplementation increased butyrate-producing bacteria, was associated with a higher incidence of aGvHD after day +100, but reduced classic aGvHD, indicating biological differences between these disease phases.^72^ We previously showed that DAT administration can modulate both GvHD and GVL via a context and dose-dependent effect, protecting from GvHD in physiological concentrations, but capable of inducing lethal, hyperacute GvHD at high doses.^67^ Similarly, a high stearic acid diet was associated with fecal acetic acid and aggravated aGvHD in mice.^73^ Moreover, a recent study reported an expansion of mucosa-derived IgA-producing plasmablasts before the onset of cGvHD, with antibodies recognizing commensal SCFA-producing bacteria, suggesting that immune reactivity against metabolite-producing taxa may contribute to cGvHD development.^74^ These findings emphasize the context-dependent roles of microbial metabolites in GvHD and the need to evaluate them across distinct post-transplant phases.

We observed that stool VA concentrations were significantly higher in patients with cGvHD (**Figure 6 D**). Emerging evidence suggests that VA can enhance immune cell proliferation, reduce T-cell exhaustion, and stimulate CAR-T cells ex vivo. ^75^ However, VA can also act as an epigenetic regulator, exerting anti-inflammatory effects via histone deacetylase inhibition^76^, a mechanism that is particularly relevant in GvHD. ^77^ Interestingly, patients with cGvHD of the skin exhibited enrichment of butyric, propionic, and VA during the post-transplant period (**Figure 6 E**). Most of the few available studies have addressed SCFAs in cGvHD without organ specificity^27^, mechanistic insights suggest that SCFAs can modulate skin barrier integrity.^78,79^ Another study reported that the presence of butyrogenic bacteria after aGVHD onset is associates with subsequent steroid-refractory GVHD or cGVHD.^80^ Similarly, in a small cohort, metagenomic pathways relevant for SCFA biosynthesis were enriched in patients developing cGvHD compared to matched controls.^32^ The lack of significant associations between *IMM-RI post-TX* and cGvHD in other organs may reflect their lower prevalence, as skin involvement was the most common in the validation cohort (skin, n = 48%; oral mucosa, n = 37%; liver n = 20%).

Lastly, we observed that early antibiotic exposure (**Table 3**) was associated with *IMM-RI postTX* high-risk, suggesting that antibiotic-driven microbiome disruption, particularly affecting microbial metabolic outputs, can persist beyond peri-engraftment and may have longer-term consequences for relapse risk and cGvHD than previously appreciated.

### Limitations

Some limitations to our study should be noted: OS observed in both cohorts was lower than established allo-HCT benchmarks, likely reflecting sampling bias toward patients with complicated post-transplant courses, including GvHD, who were sampled more frequently. As a result, complication rates may be overestimated and survival underestimated. Importantly, the significant association between IMM-RI post-TX and OS was consistently observed across both cohorts, highlighting the robustness of our findings. Nevertheless, validation in an independent, prospectively enrolled cohort with systematic sampling, such as that of the Mount Sinai Acute GVHD Consortium (MAGIC),^81^ will be essential to confirm the broader applicability of these findings.

### Outlook

No guidelines or approved therapies exist to overcome microbial dysbiosis after allo-SCT or at the onset of GvHD. Interventions such as dietary modulation, pre- or probiotics, and fecal microbiota transplantation (FMT) have been explored, with encouraging but preliminary results.^82–84^ FMT can achieve remission in up to 28-75% of patients suffering from aGvHD.^83^ Probiotics, defined as live microorganisms that confer health benefits to the host, have demonstrated promising safety profiles and beneficial effects in experimental settings.^85–87^ Prebiotics, indigestible dietary compounds such as dietary fiber, promote the production of beneficial microbial metabolites, including butyrate.^88^ However, host–microbiome interactions are complex and the relevant microbial taxa, metabolites, and mechanisms involved, as well as their long-term effects on clinical outcomes, remain insufficiently understood. Adverse effects have also been reported, including increased rates of late-onset aGvHD following prebiotic administration^72^ and even impaired efficacy of immune checkpoint inhibition associated with commercially available probiotics.^89^ An alternative approach is to directly administer protective microbial metabolites, or “postbiotics,” like butyrate or the secondary BAs, like ursodeoxycholic acid (UDCA), which have been linked to epithelial protection and improved GvHD outcomes.^18,26,90,91^ Clinical studies investigating specific metabolites in patients receiving allo-SCT are rare, and further research is needed to better characterize the microbial metabolites, especially intestinal SCFAs and BAs, in the post-transplantat period and their impact on clinical outcomes, potentially identifying therapeutic targets for long-term intervention.

Current microbiome “signatures” are defined as lists of differentially abundant taxa or taxonomy-based classifiers trained in a single cohort, yet they frequently show poor external validity when applied to independent datasets, reflecting cohort-dependence, and technical heterogeneity. Large cross-study evaluations have highlighted this limited cross-cohort reproducibility as a major challenge for microbiome biomarker research.^60^ In contrast, the *IMM-RI post-TX* emphasizes microbial metabolic output over taxonomy potentially providing more robust predictive performance across cohorts with diverse community structures shaped by geography, diet, antibiotic exposure, and technical batch effects. This approach could help clinicians stratify patients by risk and identify those requiring closer monitoring for relapse or cGvHD. Importantly, microbial metabolites are actionable via targeted interventions, such as precision FMT or direct supplementation of protective metabolites. Finally, the dual, context-dependent associations we observed may guide personalized immunotherapy strategies not only in alloHCT, but also in broader cancer immunotherapy settings, including immune checkpoint blockade and CAR-T cell therapy.

## Acknowledgments

This study was supported by the Faculty Clinician Scientist Program of the Faculty of Medicine of TUM (fellowship to A.S.), the Deutsche Forschungsgemeinschaft, –Projektnummer 395357507 – SFB 1371 (to H.P., E.T.O., E.H., E.M., K.N., K.P.J., K.K., M.S.), Projektnummer 324392634 – TRR 221 (to H.P., E.H., M.E., D.W., D.W., W.H., A.G.), Projektnummer 514894665 in TRR 387/1 (to F.B.), DFG PO 1575/5-1 (to H.P.), the German Cancer Aid (70114547 to H.P.), the Wilhelm Sander Foundation (2021.040.1 to H.P.), BA 2851/6-1 and BA 2851/7-1 to F.B., DFG under Germany’s Excellence Strategy, the Else-Kröner-Fresenius-Stiftung (funding line: Else-Kröner Forschungskolleg to E.T.O. and E.M.), the Deutsches Konsortium für Translationale Krebsforschung (fellowship to E.T.O. and A.S.), E.T.O. is supported by the DGIM Advanced Clinician Scientist Program. H.P. is supported by the EMBO Young Investigator Program and the Bavarian Cancer Research Center (BZKF) (to H.P., F.B., W.H.). This work was funded/co-funded by the European Union (project MICROBOTS, Grant No. 101124680 to H.P.). Views and opinions expressed are however those of the author(s) only and do not necessarily reflect those of the European Union or the European Research Council. Neither the European Union nor the granting authority can be held responsible for them. Schematics were generated with BioRender.

We would like to thank the REG allo-SCT team, especially H. Bremm, M. Caioni, T. Schifferstein, S. Rast and Y. Schumann for their help in collecting and cryopreserving stool samples and S. Gleich for data management. We express gratitude to the MUC allo-SCT team, especially K. Braitsch, K. Koch, L. Oßwald, K. Nickel and the entire D2a nursing staff for their sample acquisition. We acknowledge A. Conrad and W. Johannes (K.-P.J. laboratory) for help with biobanking and the MUC ColoBAC team: R. Schmid, M. Quante, M. Middelhoff, J. Horstmann and L. Fricke.

## Authorship Contributions

Conceptualization: A.S., T.E., E.T.O.

Methodology: A.S., K.N., I.L., M.P., K.K., T. Z., M. S.

Formal analysis: A.S., E.T.O., S. Holzinger, T.E.

Investigation: A.S., T.E., M.U., S. Göldel., E.T.O., E.M.,

Resources: M. V. P. H., F. B., K.-P. J., M. F., E. T. O., H. P., D. Weber., D. Wolff, E. H., M. E., W. H., A. D., S. Göttert, R. G., A. H., A. G.,

Data Curation: A.S., T.E., E.T.O.

Writing Original Draft: A.S., E.T.O

Writing Review & Editing: A.S., E.T.O.

Visualization: A.S., T.Z,

Supervision: E.T.O., S. Heidegger., K.P.J.

Project administration: A.S., T.E., E.T.O.

All authors reviewed and approved the final version of the manuscript.

## Extended Methods

### Sample Collection

For 16S rRNA gene amplicon sequencing and metagenomics, stool samples were collected in magiX PB1 microbiome preservation buffer (microBIOMix GmbH, Regensburg). For metabolite analysis, samples were either unprocessed, fresh-frozen native stool samples (inpatient samples) that were transferred to the in-house laboratory within 4 h, or stool preserved in 95% ethanol (outpatient samples). Immediately after receipt at the central laboratory (Core Facility Microbiome, ZIEL Institute for Food & Health, Technical University of Munich), sam-ples were stored at -80 °C until further processing

### 16S rRNA gene amplicon sequencing

Isolation of total DNA and sequencing of 16S rRNA gene amplicons was carried out at the Core Facility Microbiome as described previously^1^ with slight modifications. Briefly, DNA was isolated using a MaxWell (Promega, Walldorf, Germany) after bead beating. The template DNA was used in a 2-step PCR to generate sequencing libraries. The first PCR uses primers specific for the V3/V4 regions of the 16S rRNA gene (i.e., 341F, CCT ACG GGN GGC WGC AG; 785R, GAC TAC HVG GGT ATC TAA TCC) that also contain an overhang for the subsequent PCR for sample barcoding. Cleaned libraries were sequenced using PE300-cartridges v3 on a MiSeq (Illumina). For further processing of the raw amplicon sequencing data, we used IMNGS2.org. This platform is an updated version of IMNGS.org.^2^ It is based on UPARSE^3^ and UNOISE2^4^ from the USEARCH package^5^, conducting sequence quality control, chimera filtering and cluster formation of denoised amplicons (i.e., zOTU). Samples with less than 2,000 counts are removed from further analysis. Taxonomic classifications are conducted using the Taxonomy Informed Clustering (TIC) algorithm based on the database SILVA v138.1.^6^ TIC allows taxonomic placing of unknown taxa, which avoids data loss, and the cluster formed are more pure. Rhea v4.0.3 was used for downstream analysis. Firstly, zOTU tables were normalized using total sum scaling^7^ and zOTUs below a cutoff of 0.25% relative abundance were removed^8^. Next, Rhea was used for calculating alpha and beta diversity values.

### Metabolome analysis

We analyze microbial metabolites via two panels, comprising the following metabolites: SCFAs, BCFAs, IFN-I-inducing metabolites such as tryptophan derivatives, and the flavonoid metabolite DAT, lactic acid, primary and secondary BAs.

#### Sample preparation for targeted analysis

Approximately 100 mg of nativ stool was weighed in a 15-mL bead-beater tube (TeenPrepTMLysing Matrix D, MP Biomedical). In case of stool samples collected in ethanol, 400mg of the ethanol stool suspension was weighted in 15-mL bead-beater tubers. Methanol-based dehydrocholic acid extraction solvent (5 mL, c = 1.3 μmol/L) was added as an internal standard for work-up losses. Samples were extracted with a bead beater (FastPrep-24TM 5G, MP Biomedicals) supplied with a CoolPrepTM (MP Biomedicals, cooled with dry ice) three times each for 20 s with 30 s breaks in between at a speed of 6 m/s. One aliquot of the fecal suspension (1 mL) was dried in a vacuum centrifuge (Eppendorf Vacufuge) to determine the dry weight of the stool. All measured metabolite concentrations were based on the stool dry weight.

#### Measurement of SCFAs, lactic acid, desaminotyrosine and indole-3-carboxyaldehyde (metabolite panel 1)

The 3-NPH method was used for quantitation of SCFAs as well as lactic acid, DAT and indole-3-carboxyaldehyde^9^. Briefly, 40 μL of the fecal extract and 15 μL of 50 μM isotopically labeled standards were mixed with 20 μL of 120 mM EDC HCl–6% pyridine solution and 20 μL of 200 mM 3-NPH HCl solution. After 30 min at 40 °C with shaking at 1,000 rpm using an Eppendorf ThermoMixer (Eppendorf), 900 μL acetonitrile–water (50/50, v/v) was added. After centrifugation at 13,000 rpm for 2 min, the clear supernatant was used for analysis. The measurement was performed using a QTRAP 5500 triple quadrupole mass spectrometer (SCIEX) coupled to an ExionLC AD (SCIEX) ultrahigh-performance liquid chromatography system. The electrospray voltage was −4,500 V, curtain gas was at 35 psi (≈ 241 kPa), ion source gas 1 was at 55 psi (≈ 379 kPa), ion source gas 2 was at 65 psi (≈ 448 kPa), and the temperature was 500°C. Multiple-reaction-monitoring parameters were optimized using commercially available standards. Chromatographic separation was performed on a 1.7-μm, 100 × 2.1-mm, 100 Å (= 10 nm) Kinetex C18 column (Phenomenex) with 0.1% formic acid (eluent A) and 0.1% formic acid in acetonitrile (eluent B) as elution solvents. An injection volume of 1 μL and a flow rate of 0.4 mL/min were used. The gradient elution started at 23% B, which was held for 3 min; afterward the concentration was increased to 30% B at 4 min, with another increase to 40% B at 6.5 min; at 7 min, 100% B was used, which was held for 1 min; at 8.5 min, the column was equilibrated at starting conditions. The column oven was set to 40 °C, and the autosampler was set to 15 °C. Data acquisition and instrumental control were performed with Analyst v1.7 software (SCIEX). Data were analyzed with MultiQuant v3.0.3 (SCIEX) and MetaboAnalyst^10^. Features with more than 70% of missing values were removed. Missing values were replaced by limit of detection (1/5 of the minimum positive value of each variable).

#### Measurement of bile acids (metabolite panel 2)

BA analysis was performed according to Reiter et al.^11^ Briefly, 20 μL of isotopically labeled BAs (approximately 7 μM each) were added to 100 μL of fecal extract. Targeted BA measurement was performed using the same system as described above. A multiple-reaction-monitoring method was used for detection and quantification of BAs^11^. An electrospray ion voltage of −4,500 V and the following ion source parameters were used: curtain gas (35 psi ≈ 241 kPa), temperature (450 °C), gas 1 (55 psi ≈ 379 kPa), gas 2 (65 psi ≈ 448 kPa) and entrance potential (−10 V). MS parameters and LC conditions were optimized using commercially available standards of endogenous BAs and deuterated BAs for simultaneous quantification of the selected 34 analytes. For separation of analytes, a 100 × 2.1-mm, 100 Å (= 10 nm), 1.7-μm, Kinetex C18 column (Phenomenex) was used. Chromatographic separation was performed with a constant flow rate of 0.4 mL/min using a mobile phase consisting of water (eluent A) and acetonitrile–water (95/5, v/v, eluent B), both containing 5 mM ammonium acetate and 0.1% formic acid. The gradient elution started with 25% B for 2 min, increased at 3.5 min to 27% B and in 2 min to 35% B, which was held until 10 min, increased in 1 min to 43% B, held for 1 min, increased in 2 min to 58% B and held 3 min isocratically at 58% B, and then the concentration was increased to 65% at 17.5 min, with another increase to 80% B at 18 min, followed by an increase at 19 min to 100% B, which was held for 1 min. At 20.5 min, the column was equilibrated for 4.5 min at starting conditions. The injection volume for all samples was 1 μL, the column oven temperature was set to 40 °C, and the autosampler was kept at 15 °C. Data acquisition and instrumental control were performed with Analyst 1.7 software (SCIEX). Data were analyzed as indicated for panel 1.

### Metagenomics

The genomic DNA already isolated for 16S-rRNA gene amplicon sequencing, was also used here. Briefly, DNA was randomly sheared into short fragments, which were end repaired, A-tailed, and further ligated with Illumina adapters. The fragments with adapters were size-selected, PCR amplified, and purified. The library was checked with Qubit and real-time PCR for quantification and a 2100 Bioanalyzer (Agilent) for size distribution detection as required by Illumina (TruSeq® DNA Library Prep Kit, Illumina). Quantified libraries are pooled and sequenced on a NovaSeq platform (Illumina) in PE150 mode. Raw data were aimed at 5 Gbp per demultiplexed sample. The metagenomic raw reads were processed by removing adapters and trimming sequences using Trim Galore^12^ with the following parameters: --phred33 --quality 30 --stringency 5 –length 10. Then KneadData^13^ was used for quality checking, quality filtering and host sequences decontamination, by setting “--trimmomatic-options HEADCROP:15 SLIDINGWINDOW:4:15 MINLEN:50” and default parameters.

The taxonomic profiling of microbial communities was performed using MetaPhlAn v3.0^14^, to assign reads into different taxonomies and determine species relative abundance profiles, with default parameters. The functional analysis of the metagenomic samples was performed with HUMAnN 3^15^ using default parameters.

### Data availability

High-throughput sequencing and metabolomics data have been deposited in public repositories in accordance with journal guidelines. 16S rRNA gene amplicon sequencing data are available at the NCBI Gene Expression Omnibus (GEO) under project ID: PRJNA1435038 and the accession link: https://dataview.ncbi.nlm.nih.gov/object/PRJNA1435038?reviewer=eueanlm4c1frn2398cm3sdinp4.

Shotgun metagenomic data are available at the ENA database, https://www.ebi.ac.uk/ena/browser/home under the accession password: PRJEB109296.

Raw and processed targeted metabolomics data have been deposited in Massive. ftp://MSV000101054@massive-ftp.ucsd.edu under accession password: cVJAMS4xw7O41GHu. All accession links are active at the time of submission. Data will be publicly accessible upon publication.

### Statistical analyses

Alpha diversity analyses were conducted applying Rhea v4.0.3 as described above.^16^ Richness is reported as “effective richness”, i.e., number of OTUs per 1000 reads.^8^ The Shannon index was converted to effective Shannon numbers (i.e., e^Shannon^ ^index^), which transforms the index to linear numbers.^16^ Beta diversity, as a measure of similarity between different microbial profiles, was calculated using generalized UniFrac (gunif), which is less sensitive to shallow or deep parts of the taxonomic tree compared to weighted and unweighted UniFrac.^17^ Distances for every pair of samples were calculated as gunif by Rhea v4.0.3.^16^ To compare similarities between before- and post-transplantation microbiome, we calculated the percentage of initial microbiome as follows: (100-100×((gunif(day +56)+gunif(day +100))/2)). Diversity measures and metabolite concentrations were neither normally nor log-normally distributed. Data were tested for normality using the D’Agostino & Pearson, Anderson-Darling, Shapiro-Wilk, and Kolmogorov–Smirnov tests. To assess lognormality, data were log-transformed and the transformed data were subjected to the same normality tests. We applied a nonparametric Kruskal-Wallis test for multiple comparison testing for not normally distributed data, followed by Dunn’s test. The means of preselected pairs were compared: *PRE-TX* vs. *PERI-ENGRAFTMENT*, *PRE-TX* vs. *LATE POST-TX,* and *PERI-ENGRAFTMENT* vs. *LATE POST-TX*. Missing values precluded a mixed-effects analysis. For comparison testing between the time points of the *RECOVERY* and *NO RECOVERY* group, we applied repeated measures and ANOVA testing with Geisser-Greenhouse correction and used a mixed effects analysis with multiple comparison testing. Further, we corrected for multiple comparisons using Holm-Šidák test. Concentration data were analyzed using generalized linear mixed-effects models (fitted with the ‘lme4’ package in R) with a Gamma distribution and log link to account for non-normality. Disease severity was modeled either as an ordinal numeric predictor or as a binary variable (disease yes/no), time point as a categorical fixed effect, and subject as a random intercept to account for repeated measurements. Interactions between the predictor and time point were included to allow the association between severity and concentration to differ across time points. Time-point specific associations between severity and concentration were evaluated using estimated marginal trends and contrasts from the emmeans package, corresponding to Wald tests of the severity slope or contrast within each time point. For analyses involving multiple metabolites, the same model and hypothesis tests were applied independently to each metabolite. P-values for the predictor effect were adjusted for multiple testing using Benjamini-Hochberg false discovery rate (FDR). OS was estimated by the Kaplan-Meier method. Cumulative incidences for TRM, GvHD, and relapse were estimated with RStudio v2024.04.2+764^18^ using the cmprsk package and CumIncidence function.^19^ Estimates of TRM were calculated using relapse as the competing risk, and estimates for severe aGvHD were calculated using death as the competing risk. All other analyses, except for the competing risk analyses and the mixed-effects concentration models, were performed using GraphPad Prism v10. In all analyses, statistical significance was defined as p < 0.05. Schematics were generated with Biorender.

## SUPPLEMENTAL FIGURE

**Supplemental Figure 1: *IMM-RI* metabolite concentrations do not associate with effective richness values at post-TX**

**A**: Simple linear regression with 95% confidence between stool metabolite concentrations of metabolites comprising the *IMM-RI* (butyric, propionic, and isovaleric acid, DAT, and ICA) and effective richness values at post-TX. 19 patients, of whom metabolite measurements were available, were included in the analysis. P-values < 0.05 were considered significant.

**Supplemental Figure 2: Competing risk-adjusted cumulative incidence of GI-aGvHD in the *discovery cohort***

**A** Cumulative incidence of GI-aGvHD (dashed line) with death (solid line) as a competing risk between *IMM-RI post-TX* low-risk (green) and *IMM-RI post-TX* high-risk (red) group of the discovery cohort. Number of patients at risk are shown below the graph. P-values < 0.05 were considered significant. There were only 19 patients, as 1 patient had to be excluded because metabolite values were unavailable at all time points

**Supplemental Figure 3: *IMM-RI post-TX predicting***

**A**: Receiver operating characteristic curve indicating sensitivity and specificity for the *IMM-RI post-TX to* predict OS. The area under the curve (AUC), positive predictive value and negative predictive value indicated.

## SUPPLEMENTAL TABLES

**Supplemental Table 1: The metabolomic compounds of the *IMM-RI peri-engraftment* and *post-TX*.**

Metabolites denoting the *IMM-RI*. Optimized thresholds for concentrations were determined using the Youden index. A point was assigned to the metabolite in question, if concentrations were beneath the cutoff. A score ranging from 0 to 5 points could be assigned to the *IMM-RI*. Patients were categorized into *IMM-RI* lowand high-risk groups based on their scores (0 – 2 indicating *IMM-RI* low-risk and 3 – 5 indicating *IMM-RI high-risk*). Higher scores suggest microbiome damage and IMM depletion.

**Supplemental Table 2: Microbiome-related epidemiological patient characteristics in the *discovery cohort*.**

Patients are characterized as smokers if they are currently smoking or if they have ever smoked regularly in the past. Alcohol dependence is defined according to official World Health Organization (WHO) guidelines. Western diet includes the consumption of meat, regardless of frequency. Physical activity is defined as regular exercise of at least 1 hour per week. Body weight is classified using the body-mass index (BMI) and assigned to pre-defined BMI categories.

**Supplemental Table 3: Beginning of antibiotic treatment according to *RECOVERY status in discovery cohort*.**

Distribution of BSA administration, including piperacillin-tazobactam and carbapenems, by timing (*PRE-TX* and *PERI-ENGRAFTMENT*) and by RECOVERY and NO RECOVERY. BSA administration that started in the *PRE-TX* time point and continued into *PERI-ENGRAFTMENT* was counted only in *PRE-TX*.

**Supplemental Table 4: Immunosuppressive therapy after allo-SCT.**

Values exceeding 100 arise because immunosuppressive therapies were administered in combination and, in some cases, substituted. MMF, mycophenolate mofetil; ATG, anti-thymocyte globulin.

**Supplemental Table 5: Microbial metabolites quantified by targeted mass spectrometry.** The major metabolite classes are shown, including SCFAs, BCFAs, primary BAs, secondary BAs, Flavonoids and Indoles. For each class, the individual metabolites are listed, whose concentrations in stool samples were quantified by targeted mass spectrometry. Only metabolites included in the statistical analyses are shown.

**Supplemental Table 6: Distribution of similarity between the *RECOVERY* and *NO RECOVERY*.**

Similarity percentages between *POST-TX* and *PRE-TX* are indicated for every patient. Patients with similarity values above 52.45 were stratified into “*SIMILARITY*”, otherwise as “*NO SIMILARITY*”.

**Supplemental Table 7: Pathways included in the butanoate superclass pathway.**

Pathways assigned to the butanoate superclass according to the MetaCyc database

