## Supplemental Figures 1-3 and Tables 1-7 for "Immunomodulatory metabolites define long-term gut microbiome recovery after allogeneic HCT and associate with improved survival and reduced relapse related mortality"

### Supplemental Figure 1

A

***IMM-RI* metabolites according to effective richness at *POST-TX***

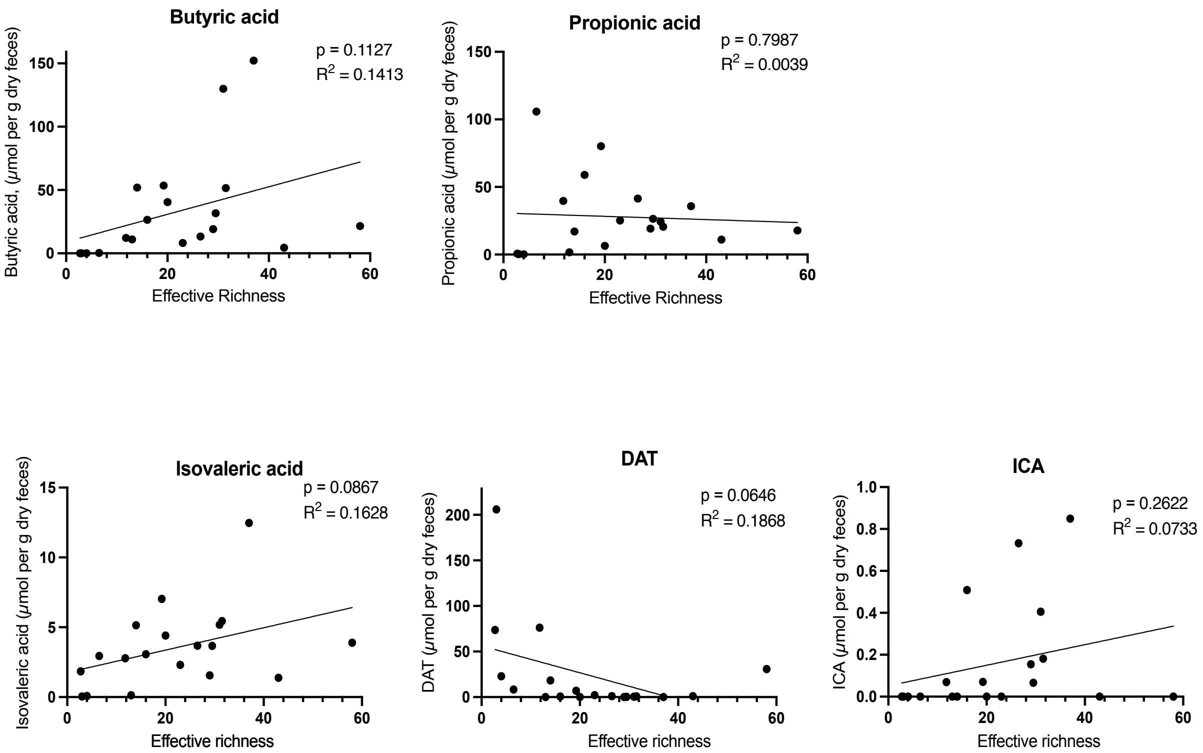

### Supplemental Figure 2

A

**Incidence of GI-aGvHD / death**

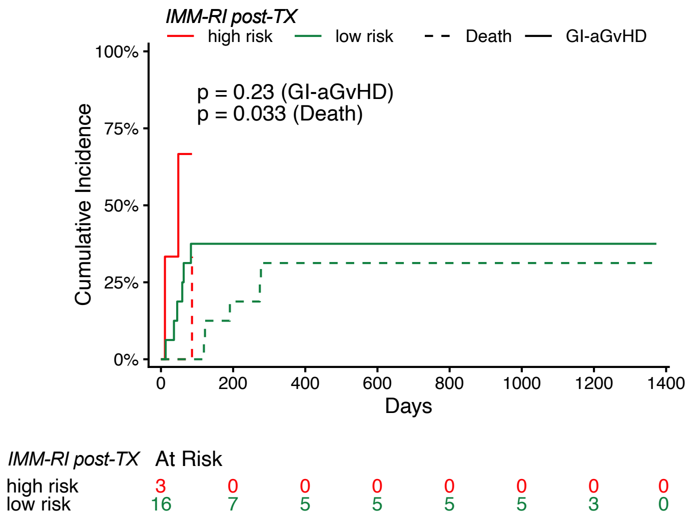

### Supplemental Figure 3

A

***IMM-RI post-TX* low-risk predicting 2-year OS**

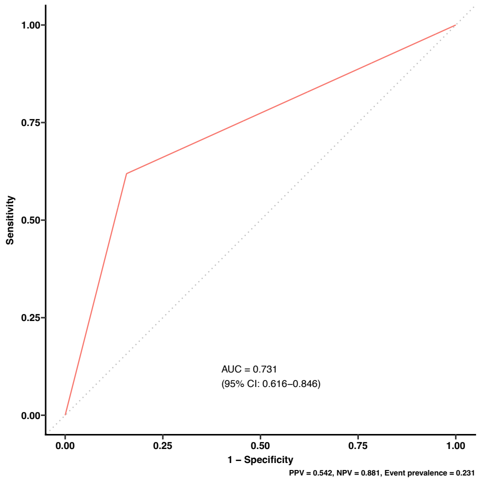

**Supplemental Table 1. The metabolomic compounds of the *IMM-RI peri-engraftment* and *IMM-RI post-TX***

| Compound *IMM-RI peri-engraftment* | 0 points | 1 point |
|---|---|---|
| Propionic acid [µmol/g dry feces] | > 21.96 | ≤ 21.96 |
| Butyric acid [µmol/g dry feces] | > 1.41 | ≤ 1.41 |
| Isovaleric acid [µmol/g dry feces] | > 0.035 | ≤ 0.035 |
| Desaminotyrosine [µmol/g dry feces] | > 0.005 | ≤ 0.005 |
| Indolecarboxyaldehyde [µmol/g dry feces] | > 0.035 | ≤ 0.035 |

| Compound *IMM-RI post-TX* | 0 points | 1 point |
|---|---|---|
| Propionic acid [µmol/g dry feces] | > 19.15 | ≤ 19.15 |
| Butyric acid [µmol/g dry feces] | > 4.052 | ≤ 4.052 |
| Isovaleric acid [µmol/g dry feces] | > 4.156 | ≤ 4.156 |
| Desaminotyrosine [µmol/g dry feces] | > 0.1338 | ≤ 0.1338 |
| Indolecarboxyaldehyde [µmol/g dry feces] | > 0.0017 | ≤ 0.0017 |

**Supplemental Table 3. Beginning of antibiotic treatment according to *RECOVERY* status in *discovery* cohort**

| AB timing - no. (%) | *RECOVERY* n = 9 | *NO RECOVERY* n = 11 |
|---|---|---|
| **No BSA** | 0 | 0 |
| **BSA in *PRE-TX*** | 1 (13) | 2 (18) |
| **BSA in *PERI-ENGRAFTMENT*** | 8 (87) | 9 (82) |

#### Supplemental Table 4. Immunosuppressive therapy after allo-SCT

| **Immunosuppressive Therapy - no (%)** | ***RECOVERY* n = 9** | ***NO RECOVERY* n = 11** |
|---|---|---|
| ATG | 9 (100) | 8 (73) |
| MMF | 9 (100) | 11 (100) |
| Cyclosporin A | 7 (16) | 6 (55) |
| Tacrolimus | 2 (22) | 6 (55) |
| post-transplant Cyclophosphamide | 0 ( 0) | 3 (27) |
| Sirolimus | 0 ( 0) | 1 (9) |

**Supplemental Table 5. Microbial metabolites quantified by targeted mass spectrometry**

| Class | Metabolite |
|---|---|
| **SCFAs** | Acetic acid |
| | Butyric acid |
| | Propionic acid |
| | Valeric acid |
| **BCFAs** | Isobutyric acid |
| | 2-Methylbutyric acid |
| | Isovaleric acid |
| **Primary BAs** | Cholic acid |
| | Chenodeoxycholic acid |
| | Glycocholic acid |
| | Glycochenodeoxycholic acid |
| | Taurochenodeoxycholic acid |
| **Secondary BAs** | Dehydrocholic acid |
| | Glycodeoxycholic acid |
| | Lithocholic acid |
| | Ursodeoxycholic acid |
| | Deoxycholic acid |
| | Taurodeoxycholic acid |
| | Tauroursodeoxycholic acid |
| | Glycoursodeoxycholic acid |
| | Ursocholic acid |
| | Dehydrolithocholic acid |
| | Glycohyocholic acid |
| | 3-Dehydrocholic acid |
| | 6-Ketolithocholic acid |
| | 7-Dehydrocholic acid |
| | Cholic acid-7ol-3one |
| **Flavonoids** | Desaminotyrosine |
| **Indoles** | Indole-3-carboxyaldehyde |
| | Indole-3-acetate |

#### Supplemental Table 6. Distribution of similarity between *RECOVERY* and *NO RECOVERY*

| *RECOVERY* | *SIMILARITY (%)* | *NO RECOVERY* | *SIMILARITY (%)* |
|---|---|---|---|
| **PAT B** | 56 | **PAT A** | 33 |
| **PAT D** | 67 | **PAT C** | 45 |
| **PAT E** | 70 | **PAT F** | 93 |
| **PAT I** | 46 | **PAT G** | 29 |
| **PAT K** | 53 | **PAT H** | 30 |
| **PAT O** | 46 | **PAT J** | 32 |
| **PAT P** | 73 | **PAT L** | 48 |
| **PAT R** | 36 | **PAT M** | 54 |
| **PAT S** | 57 | **PAT N** | 52 |
| | | **PAT Q** | 69 |
| | | **PAT T** | 61 |

#### Supplemental Table 6. Distribution of similarity between *RECOVERY* and *NO RECOVERY*

| *RECOVERY* | *SIMILARITY* (%) | *NO RECOVERY* | *SIMILARITY* (%) |
|---|---|---|---|
| **PAT B** | 56 | **PAT A** | 33 |
| **PAT D** | 67 | **PAT C** | 45 |
| **PAT E** | 70 | **PAT F** | 93 |
| **PAT I** | 46 | **PAT G** | 29 |
| **PAT K** | 53 | **PAT H** | 30 |
| **PAT O** | 46 | **PAT J** | 32 |
| **PAT P** | 73 | **PAT L** | 48 |
| **PAT R** | 36 | **PAT M** | 54 |
| **PAT S** | 57 | **PAT N** | 52 |
| | | **PAT Q** | 69 |
| | | **PAT T** | 61 |

**Supplemental Table 7. Pathways included in the butanoate superclass pathway**

| Butanoate Superclass Pathway |
|---|
| PWY_5022 |
| PWY_5676 |
| P162_PWY |
| GLUDEG_II_PWY |
| PWY_8190 |
| P163_PWY |
| CENTFERM_PWY |
| PWY_5677 |

#### Supplemental Table 2. Microbiome-related epidemiological patient characteristics

| **Total patients - no. (%)** | **20 (100)** | ***RECOVERY* n = 9 (45)** | ***NO RECOVERY* n = 11 (55)** | **p-value** |
|---|---|---|---|---|
| **Consumption of stimulants - no. (%)** | | | | |
| Smoking | 14 (70) | 7 (78) | 7 (64) | 0.6424 |
| Alcohol dependance | 5 (25) | 1 (11) | 4 (36) | 0.3383 |
| **Diet - no. (%)** | | | | |
| Western diet | 20 (100) | 9 (100) | 11 (100) | >0.9999 |
| Other | 0 (0) | 0 (0) | 0 (0) | |
| **Physical activity - no. (%)** | 15 (75) | 6 (67) | 9 (81) | 0.6169 |
| **Body weight - no. (%)** | | | | |
| Normal weight (18,5 - 24,9 kg/m2) | 10 (50) | 5 (56) | 5 (45) | >0.9999 |
| Overweight (25 - 29,9 kg/m2) | 7 (35) | 3 (33) | 4 (36) | >0.9999 |
| Obesity (≥ 30 kg/m2) | 3 (15) | 1 (11) | 2 (18) | >0.9999 |
